# Physician-level classification performance across multiple imaging domains with a diagnostic medical foundation model and a large dataset of annotated medical images

**DOI:** 10.1101/2025.05.30.25328646

**Authors:** Alexander Henry Thieme, Tahir Miri, Alexandre R Marra, Takaaki Kobayashi, Guillermo Rodriguez-Nava, Yiheng Li, Thomas Barba, Ahmet Görkem Er, Justus Benzler, Maximilian Gertler, Mareike Riechers, Christian Hinze, Yuanning Zheng, Konstantin Pelz, Divya Nagaraj, Alexa Chen, Anastassia Löser, Alexander Rühle, Constantinos Zamboglou, Lujain Alyahya, Maximilian Uhlig, Gautam Machiraju, Kuba Weimann, Christoph Lippert, Tim Conrad, Jackie Ma, Roberto Novoa, Michael Moor, Tina Hernandez-Boussard, Mohammed Alawad, Jorge L. Salinas, Mirja Mittermaier, Olivier Gevaert

## Abstract

A diagnostic medical foundation model (MedFM) is an artificial intelligence (AI) system engineered to accurately determine diagnoses across various medical imaging modalities and specialties. To train MedFM, we created the PubMed Central Medical Images Dataset (PMCMID), the largest annotated medical image dataset to date, comprising 16,126,659 images from 3,021,780 medical publications. Using AI- and ontology-based methods, we identified 4,482,237 medical images (e.g., clinical photos, X-rays, ultrasounds) and generated comprehensive annotations. To optimize MedFM’s performance and assess biases, 13,266 images were manually annotated to establish a multimodal benchmark. MedFM achieved physician-level performance in diagnosis tasks spanning radiology, dermatology, and infectious diseases without requiring specific training. Additionally, we developed the Image2Paper app, allowing clinicians to upload medical images and retrieve relevant literature. The correct diagnoses appeared within the top ten results in 88.4% and at least one relevant differential diagnosis in 93.0%. MedFM and PMCMID were made publicly available.

**Funding:** Research reported here was partially supported by the National Cancer Institute (NCI) (R01 CA260271), the Saudi Company for Artificial Intelligence (SCAI) Authority, and the German Federal Ministry for Economic Affairs and Climate Action (BMWK) under the project DAKI-FWS (01MK21009E). The content is solely the responsibility of the authors and does not necessarily represent the official views of the National Institutes of Health.

## Introduction

The application of artificial intelligence (AI) to medical imaging is transforming the medical field, demonstrating potential in early detection^1–5^, diagnosis^6–24^, and treatment^7,25–28^ of diseases. However, the success of medical AI relies heavily on the quantity, quality, and diversity of training data.

The availability of ImageNet^29^, a large, manually annotated dataset containing approximately 14 million images of everyday objects, was pivotal in advancing AI. Medical images, however, are fundamentally different, as they contain pixel-level data critical for diagnosing, treating, and studying diseases. These images span diverse modalities, including clinical photography, digital histopathology, endoscopy, ultrasound, X-ray, computed tomography (CT), magnetic resonance imaging (MRI), and other imaging technologies, each requiring specialized interpretation. We proposed that a similarly extensive dataset of annotated medical images combined with a medical model pre-trained on this dataset could significantly reduce the data needed to create medical AI models for specific purposes and enhance robustness across diverse medical applications.

Single categories were inadequate for capturing the full information within a medical image, as multiple dimensions - such as diagnoses, symptoms, diagnostic tests, and treatments - were essential for informed medical decision-making^30^. Consequently, medical images required annotation with comprehensive medical text, and AI techniques were needed to interpret this detailed information. While large language models (LLMs) had been effectively applied to extensive medical text corpora^31–34^, the development of a generalist medical foundation model (MedFM) capable of integrating both visual and textual data across all medical disciplines remained unexplored. MedFM enables holistic decision-making by combining multimodal data for comprehensive diagnostics, enhances accuracy through cross-validation, and accelerates diagnosis and treatment by processing large datasets in real time. The model fosters personalized care, interdisciplinary collaboration, and standardization, making it highly scalable and adaptable across specialties and resource settings. It supports medical education, research, and innovation by identifying patterns and insights across disciplines. By unifying medical data, MedFM streamlines workflows, optimizes resources, and may improve patient outcomes.

Therefore, we firstly built the largest publicly available dataset of multimodal medical images, comparable in scale to ImageNet, by identifying and extracting medical images and associated text from the PubMed Central (PMC), namely the PMC Medical Image Dataset (PMCMID). We augmented figure captions with medical concepts extracted from the publication text using a fine-tuned LLM and a Unified Medical Language System (UMLS)^35^ parser. Secondly, we created a multimodal, manually annotated benchmark (n=13,266 PMCID images) to evaluate MedFM’s performance across different modalities, age groups, sexes, and skin tones. Thirdly, we trained MedFM using an open-source implementation of the Contrastive Language–Image Pre-training (CLIP) architecture (OpenCLIP^36^) capable of visual and text comprehension and optimized its performance using our benchmark and supercomputing resources provided by Juwels Booster^37^. These steps are automated in a pipeline for visual-language knowledge extraction from PMC **(Fig. 1A**), enabling periodic updates to PMCMID and MedFM as PMC content evolves.

**Fig. 1:**
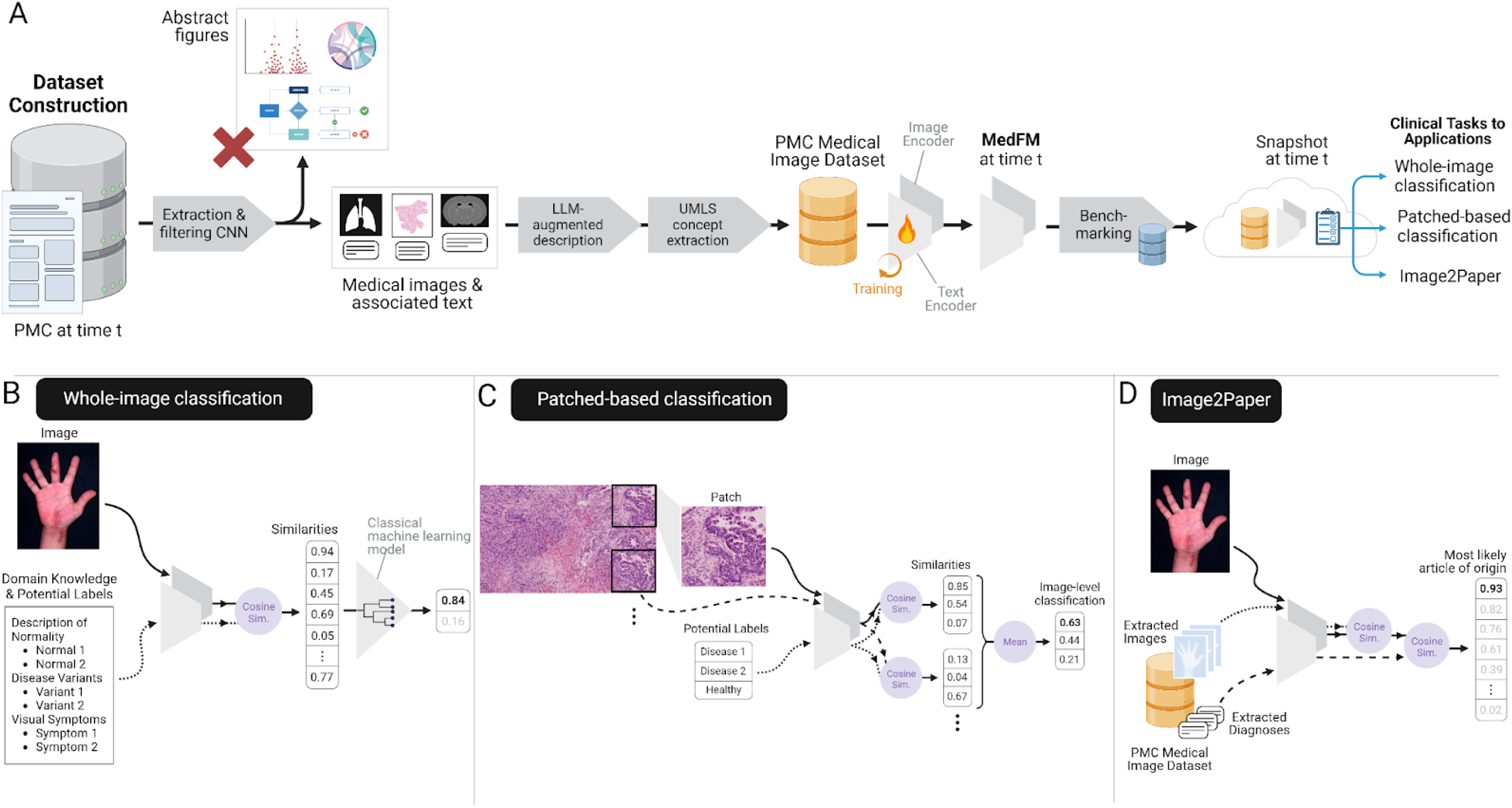
PMCMID and the MedFM pipeline for visual knowledge extraction. **A**: This panel illustrates the automated pipeline for extracting and utilizing visual-language knowledge from PubMed Central (PMC) scientific publications. The process begins with the download of all Open Access publications from PMC. A CNN then filters out non-medical images, such as abstract drawings and graphs. Medical images are augmented with comprehensive descriptions generated from the publication using a fine-tuned Stanford BioMedLM and a UMLS parser to extract medical concepts relevant for medical decision-making. These enriched medical images from the PMC Medical Images Dataset at time t (PMCMID_t). The Medical Foundation Model (MedFM_t) is trained on this dataset and evaluated using the PMCMID Benchmark, which includes over 13,000 physician-annotated medical images. MedFM_t supports various clinical tasks, such as classification and Image2Paper. With continual updates in the visual language knowledge, this pipeline periodically generates improved versions, PMCMID_t+1 and MedFM_t+1. **B:** Whole-image classification: Cosine similarities between the input image and medically relevant terms are computed by MedFM. These similarities are used as inputs to a classical machine learning model (e.g., random forest) trained on a select dataset (50-200 images) to refine MedFM’s diagnostic predictions. **C:** Patch-based classification: For high-resolution images, such as histopathology slides, the image is segmented into smaller patches. MedFM computes zero-shot classification for each patch, and the aggregated predictions (e.g., mean of all predictions) are utilized for final image classification. **D**: Image2Paper app: Users can upload a medical image to receive matches from the PMCMID dataset. MedFM first identifies the most similar images by comparing image embeddings and then ranks these images based on cosine similarities with LLM-extracted diagnoses.

We evaluated MedFM’s ability to classify diagnoses (**Fig. 1B, C**) in clinical datasets without additional training (zero-shot performance) and when combined with a classical machine learning model (CMLM) trained on limited domain-specific images. The evaluation covered prevalent diseases with major health impacts - pneumonia (radiology), skin cancer (dermatology), and lung cancer (histopathology) - as well as mpox, a rare but emerging infectious disease.

Additionally, we assessed MedFM’s capacity for improvement over time through retrospective PMCMID evaluations. To showcase its versatility, we developed the ‘Image2Paper’ app prototype (**Fig. 1D**), enabling clinicians to input a medical image and retrieve similar images with publication details from PMCMID.

## RESULTS

### Construction of the Pubmed Central Medical Image Dataset (PMCMID), description of size, image modalities, and disciplines

To create PMCMID, we extracted 16,126,659 images from 3,021,780 open access PMC articles (up to January 30, 2023). A convolutional neural network (CNN) (**Supplementary Fig. 1**) classified images into four types: Type I (individual medical images, n=656,818), Type II (image panels, n=1,745,444), Type III (mixed panels with medical images and abstract visuals, n=2,080,065), and Type IV (abstract visuals only, n=11,644,332; **Extended Data Table 1**).

To study scaling effects, we generated annual PMCMID datasets (2010–2023), growing from 504,362 images in 2010 to 16,104,074 in 2023 (**Extended Data Table 1**). Validation (n=6,730) and test (n=6,536) cohorts were created by randomly sampling 1% of Type I images, while the remaining 98% of Type I and all Type II and III images were used for training.

### Automated Annotation of PMCMID Training Cohort

PMCMID figure captions lacked key medical concepts, with diagnoses missing in 78.1% of the PMC validation set. We developed automated annotation algorithms to enrich captions into comprehensive image descriptions in the training cohort: 1) We fine-tuned BioMedLM^38^, to extract diagnoses from the associated texts (paper title, referring paragraph to the image); 2) we extracted medical concepts and UMLS Concept Identifiers (CUIs) belonging to 13 UMLS semantic types from sentence(s) and paragraph(s) referring to the image, and title of the paper. We extracted 2,158,499 unique diagnoses and 60,023,703 medical concepts (95,898 unique CUIs) from the referring sentences, 211,425,969 medical concepts (137,197 unique CUIs) from the referring paragraphs, and 16,046,845 medical concepts (68,943 unique CUIs) from the publication title.

### Manual Annotation of the PMCMID validation and test cohorts and creation of a Benchmark

The validation and test sets (n=13,266 Type I images) were annotated by 17 physicians using standardized labeling (**Supplementary Methods:** Labeling instructions). Categories included image description, diagnosis, sex, skin tone, modality, body region, and specialty (**Extended Data Table 2**). After cleaning, the validation set comprised 4,470 images and the test set 4,268. We established a benchmark using the Recall at N (R@N) metric to assess MedFM’s performance, identifying biases across sub-cohorts. MedFM achieved an R@1 of 0.65 across all images (**Fig. 2B, Supplementary Table 1**) but performed lower for patients with darker skin tones (Fitzpatrick V–VI) and females (p<0.001, **Fig. 2A, B, Supplementary Table 1**). Among body regions, head, neck, and abdomen had the highest R@1, while photographs and CTs outperformed other modalities, with microscopy performing worst (p<0.001).

**Fig 2.**
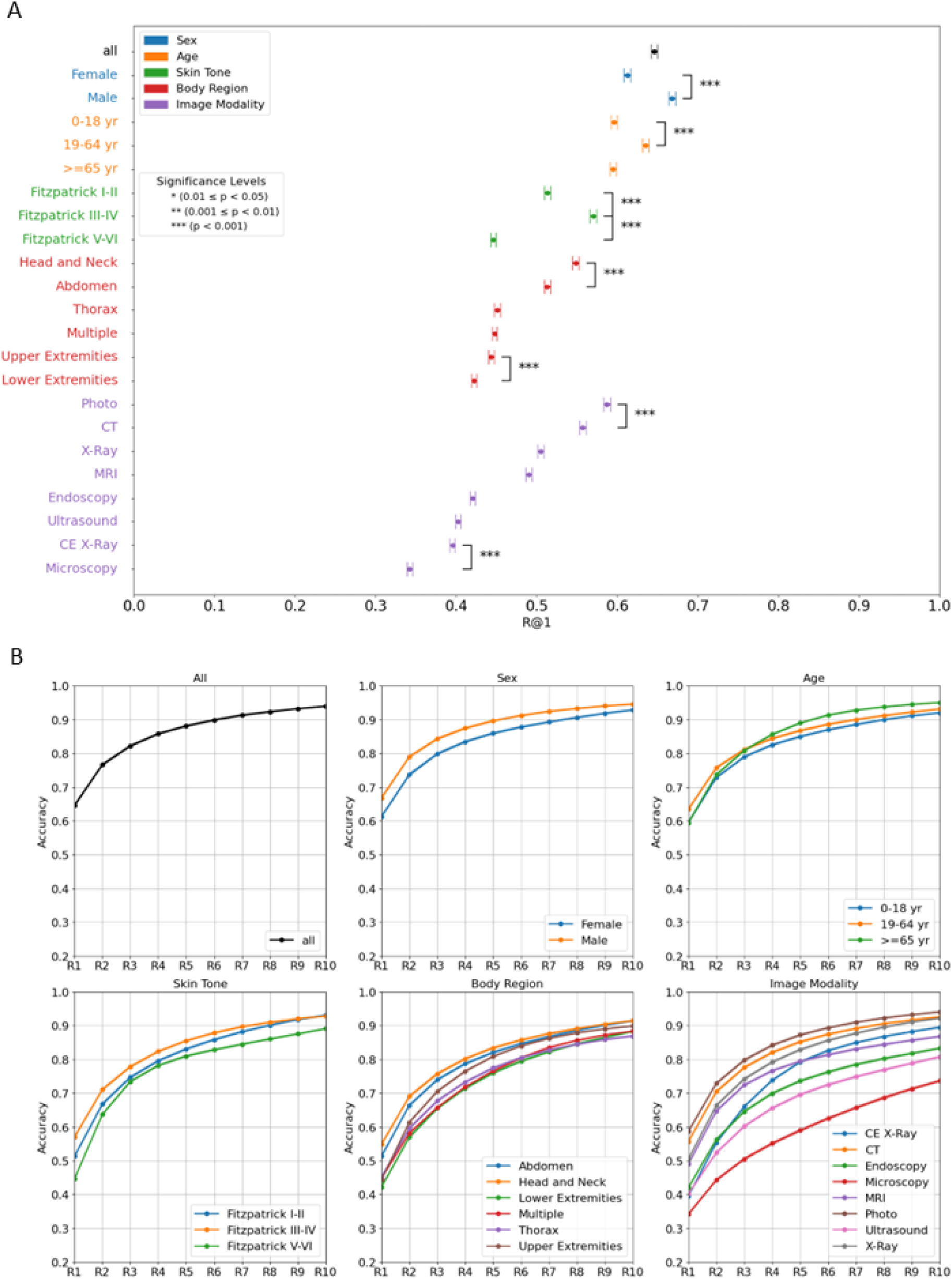
PMCMID Benchmark. Physician-annotated medical images are the basis of the multimodal PMCMID Benchmark. The benchmark repeatedly selected 50 random medical images from sub-cohorts (e.g., male and female patients) and measured the ability of the MedFM to correctly assign the diagnosis to each image. The benchmark was intended to measure performance differences between patient groups, including vulnerable groups like children or patients with darker skin tones. The benchmark used the recall at rank N (R@N) metric. R@1 reflected the percentage of images with correctly assigned diagnoses, while R@N captured the percentage of images where the diagnosis was ranked within the top N positions. To ensure reliability, the process was repeated 1000 times per subcohort to calculate average R@N and 95% confidence intervals (CIs). R@1 plots reveal statistically significant differences between patient groups, for example, between female and male patients (p < 0.001), adult patients (19-64 yr) vs children (0-18 yr) and elderly (>= 65 yr), or patients with skin tones III-IV vs. Fitzpatrick V-VI or Fitzpatrick I-II (p < 0.001). Cumulative rank plots detail R@1 till R@10 performances.

While R@1 was the primary metric, as top-ranked accuracy was critical in medicine, examining higher ranks (e.g., R@2 to R@10) could be valuable in complex clinical scenarios. To assess this, we analyzed cumulative rank plots to evaluate MedFM’s performance from R@1 to R@10. An accuracy higher than 0.9 was achieved at R@6 in all images (**Fig. 2D**). Age groups showed no differences at R@1, but adults and children performed significantly worse than the elderly starting at R@4 (**Fig. 2D**).

We analyzed MedFM’s scaling effects (**Extended Fig. 2A–F**), finding a strong correlation between R@1 performance and the logarithm of the number of training samples (R²: 0.90–0.99). Performance disparities emerged among subgroups: Fitzpatrick V–VI skin tones consistently underperformed compared to lighter tones, with projections indicating widening gaps (**Extended Fig. 2D**). Gender differences were also observed (**Extended Fig. 2F**), though performance improvements were similar.

We compared MedFM to BiomedCLIP (**Supplementary Fig. 2**), with MedFM achieving significantly higher performance across all images (R@1 = 0.65 vs. 0.59; improvement 0.056; p < 0.001) and in 16 of 22 sub-cohorts (mean R@1 improvement = 0.053; range 0.016–0.127; p < 0.001).

### Interactive UMAPs for MedFM’s image and text embeddings

We used Uniform Manifold Approximation and Projection (UMAP)^39^ to visualize image and text embeddings of the validation set (**Fig. 3; Supplementary Figs. 3-7**), highlighting relationships and clustering patterns across categories (body region, modality, and specialty).

**Fig 3.**
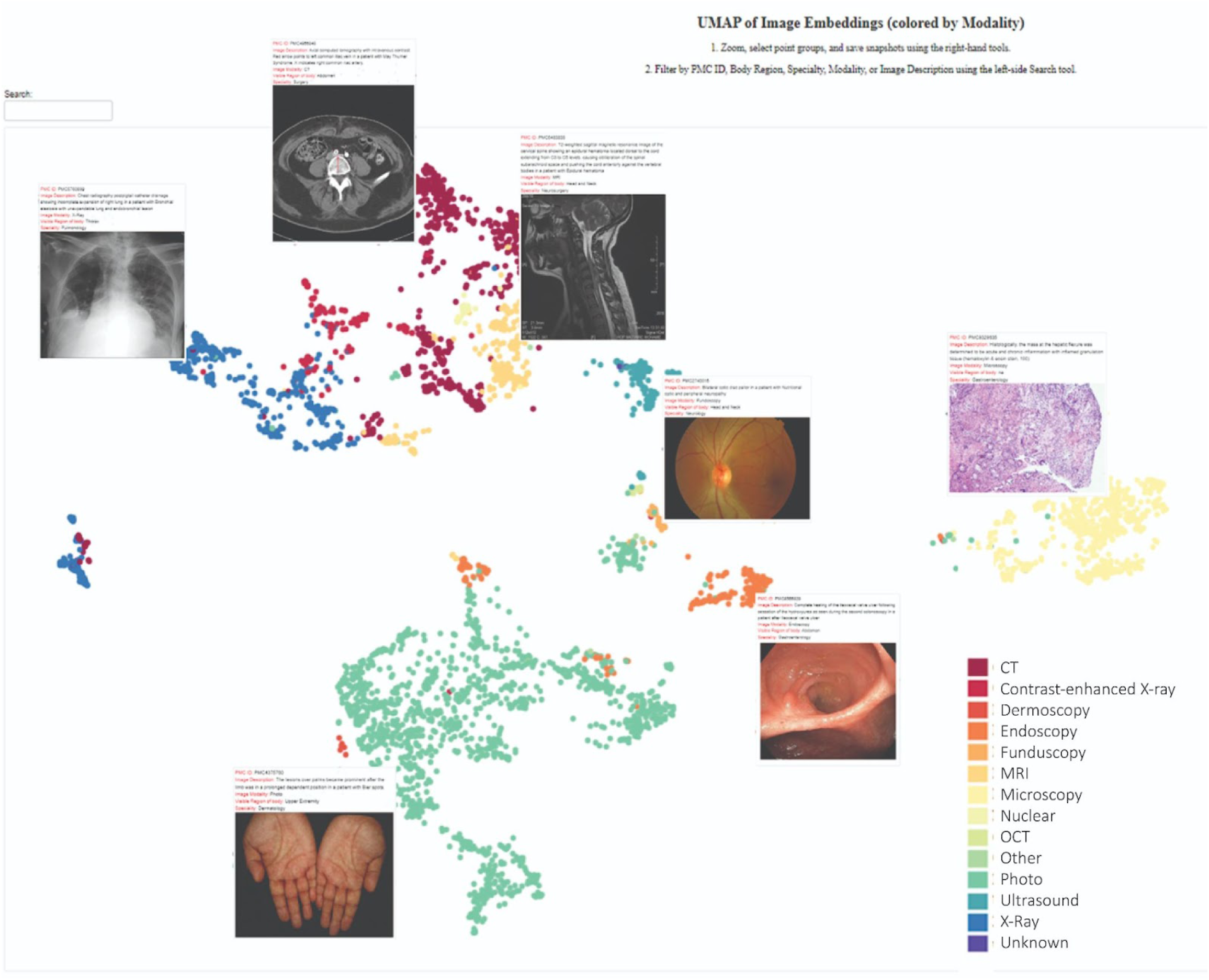
Screenshot of Interactive UMAP Visualization of the MedFM Image Embeddings. UMAP projection of image embeddings from the PMCMID Validation cohort, color-coded by the image modality, e.g. photo, CT, highlighted clustering patterns to correlate with image modalities. Selected examples from each cluster are showcased. Further UMAP analysis revealed that clusters are also aligned for anatomical body regions and medical specialities, both in the embedding space of images and texts of the MedFM. All interactive UMAPs for image and text embeddings are available online.

Image embedding clusters color-coded by modality (**Fig. 3**) showed distinct groups for photos, microscopy, and radiology, indicating MedFM captured modality relationships. Further analysis revealed modality-specific micro-clusters by anatomical region (e.g., head, thorax). Proximity between CT, MRI, and X-ray clusters of the same region suggested MedFM’s understanding of anatomical relationships. Eye images from different modalities (photos, fundoscopy, OCT) also formed closely related clusters.

Clusters emerged for medical specialties such as dermatology, dentistry, and ophthalmology (**Supplementary Fig. 4**). UMAP visualizations of text embeddings (**Supplementary Figs. 5 - 7**) showed less distinct clustering than image embeddings but still reflected relationships by modality, body region, and specialty. Interactive UMAP visualizations of MedFM’s embeddings are available online^40^.

### Image-Text Explainability

To interpret MedFM’s predictions, SHapley Additive exPlanations (SHAP) values were calculated for visual and textual inputs, highlighting feature contributions and showing strong alignment between important regions and relevant visual or textual features (**Fig. 4**).

**Fig 4:**
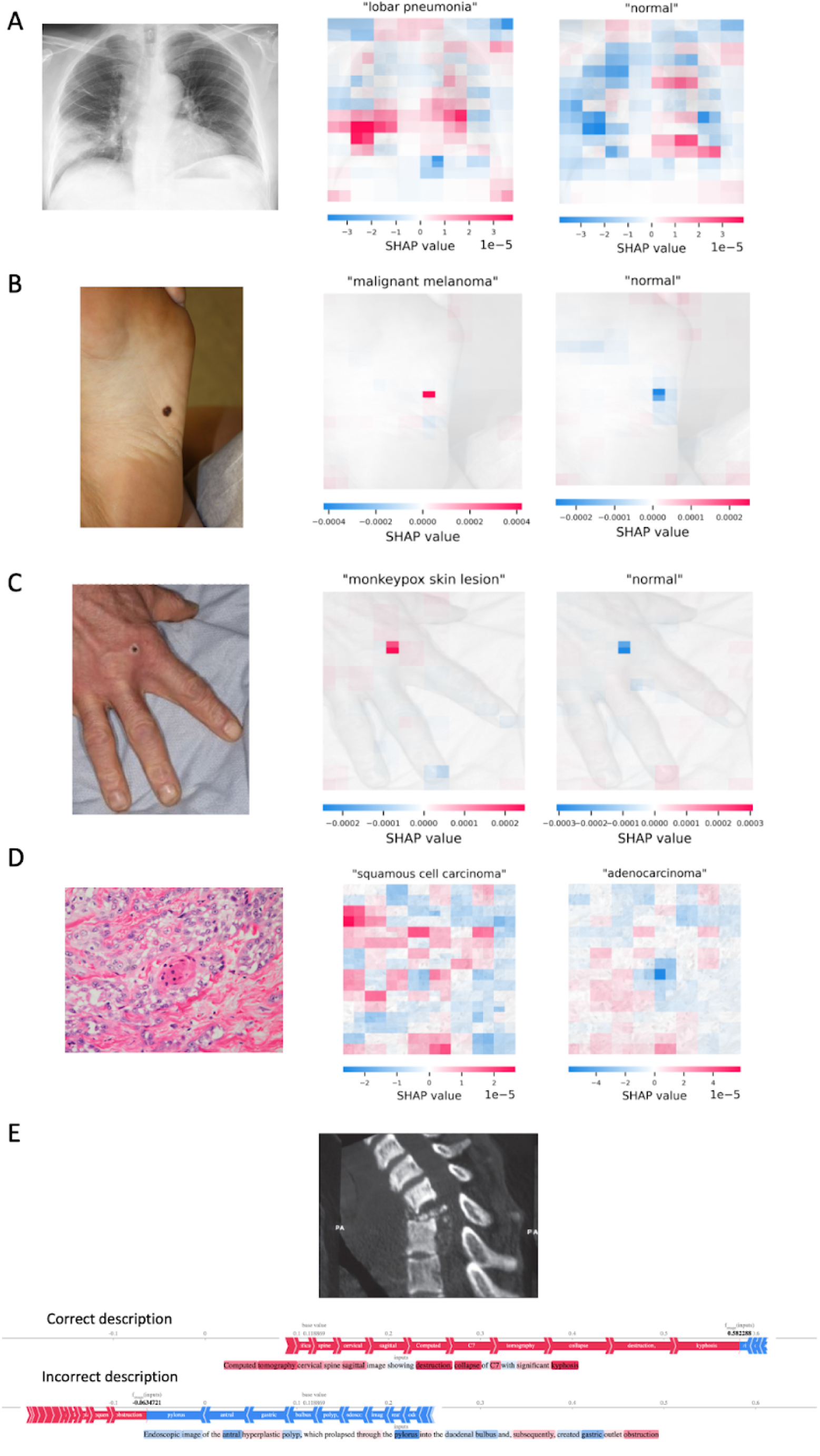
SHAP Analyses Across Multiple Medical Imaging Modalities. This figure illustrated how discriminative regions in images and key words in textual inputs were identified by SHAP analysis. A) a chest X-ray depicting lobar pneumonia in the right middle lobe; B) a clinical photo showing malignant melanoma on a foot; C) a clinical photo displaying an mpox lesion on a hand; D) a histopathology slide of squamous cell carcinoma. The left column of A-D featured medical images from various modalities, including CT, clinical photography, and histopathology. Corresponding SHAP overlays were shown in the middle and right columns. The middle overlays displayed SHAP values associated with the correct diagnosis, while the right overlays demonstrated values for an incorrect prompt, e.g., “normal”. Positive SHAP values were highlighted in red, indicating regions that increased the prediction, whereas negative values were shown in blue, indicating regions that decreased the prediction. Notably, the same regions were highlighted in both SHAP analyses: in red when contributing positively to the correct diagnosis and in blue when detracting from the prediction of wrong image description (”normal” or an incorrect diagnosis). E presented a CT image of a C7 vertebral collapse, with the upper SHAP analysis for the correct image description and the lower SHAP analysis for an incorrect description.

SHAP analyses revealed consistent discriminative regions across imaging modalities. In a chest X-ray of lobar pneumonia, regions associated with “lobar pneumonia” were highlighted in red (**Fig. 4A, middle column**), indicating positive contributions, while the same regions appeared in blue when prompted with “normal,” detracting from predicting normalcy (**Fig. 4A, right column**). Similar patterns were observed in clinical photos of malignant melanoma (**Fig. 4B**) and a mpox skin lesion (**Fig. 4C**). In a histopathology slide of squamous cell carcinoma, red regions corresponded to “squamous cell carcinoma,” while prompting with “adenocarcinoma” highlighted a blue region showing keratinization, a feature specific to squamous cell carcinoma (**Fig. 4D**).

For text inputs, SHAP values emphasized the importance of diagnostic terms in predictions. In a CT scan of spine destruction (**Fig. 4E**), terms like “destruction,” “collapse,” and “kyphosis” had the highest SHAP values, positively influencing the prediction. In contrast, mismatched descriptions had negative SHAP values, detracting from the prediction and highlighting their incompatibility with the image. These results demonstrate MedFM’s ability to align key diagnostic terms with image content, effectively identifying relevant visual and textual features for accurate predictions.

### Performance of the MedFM in four different clinical classification tasks: Detection of pneumonia in chest X-ray (CRX) images

Pneumonia is the leading infectious cause of death globally, with chest X-rays (CRXs) routinely used for diagnosis^41^. MedFM’s performance was tested using CRX images with physician-annotations across four datasets, identifying 5,885; 26,432; 2,004; and 30,227 images in CheXpert^42^, MIMIC-CRX^43^, PadChest^44^, and RSNA^45^, respectively, and a total of 64,548 CRX images (**Supplementary Table 2**). We identified 26 pneumonia classification terms (**Supplementary Table 3**).

MedFM_2023 demonstrated a zero-shot performance of AUC=0.822 and a balanced accuracy of 0.761, matching physician-level performance at AUC=0.823 (95% CI: 0.779-0.856) (**Fig. 5A**, **Extended Fig. 1A, Supplementary Table 4**). Performance increased by combining MedFM’s predictions with a CMLM, which was trained on domain-specific images from the CheXpert (n=5,885) dataset separate from the PMCMID (**Fig. 1B**). The CMLM achieved AUCs of 0.883 (95% CI: 0.879-0.887), 0.871 (95% CI: 0.860-0.881), 0.887 (95% CI: 0.881-0.892), and 0.890 for 50, 100, 200, and all available domain-specific images (n=5,885), respectively (**Extended Fig. 1A**). Physician-level performance was exceeded starting at 50 domain-specific images.

**Fig 5:**
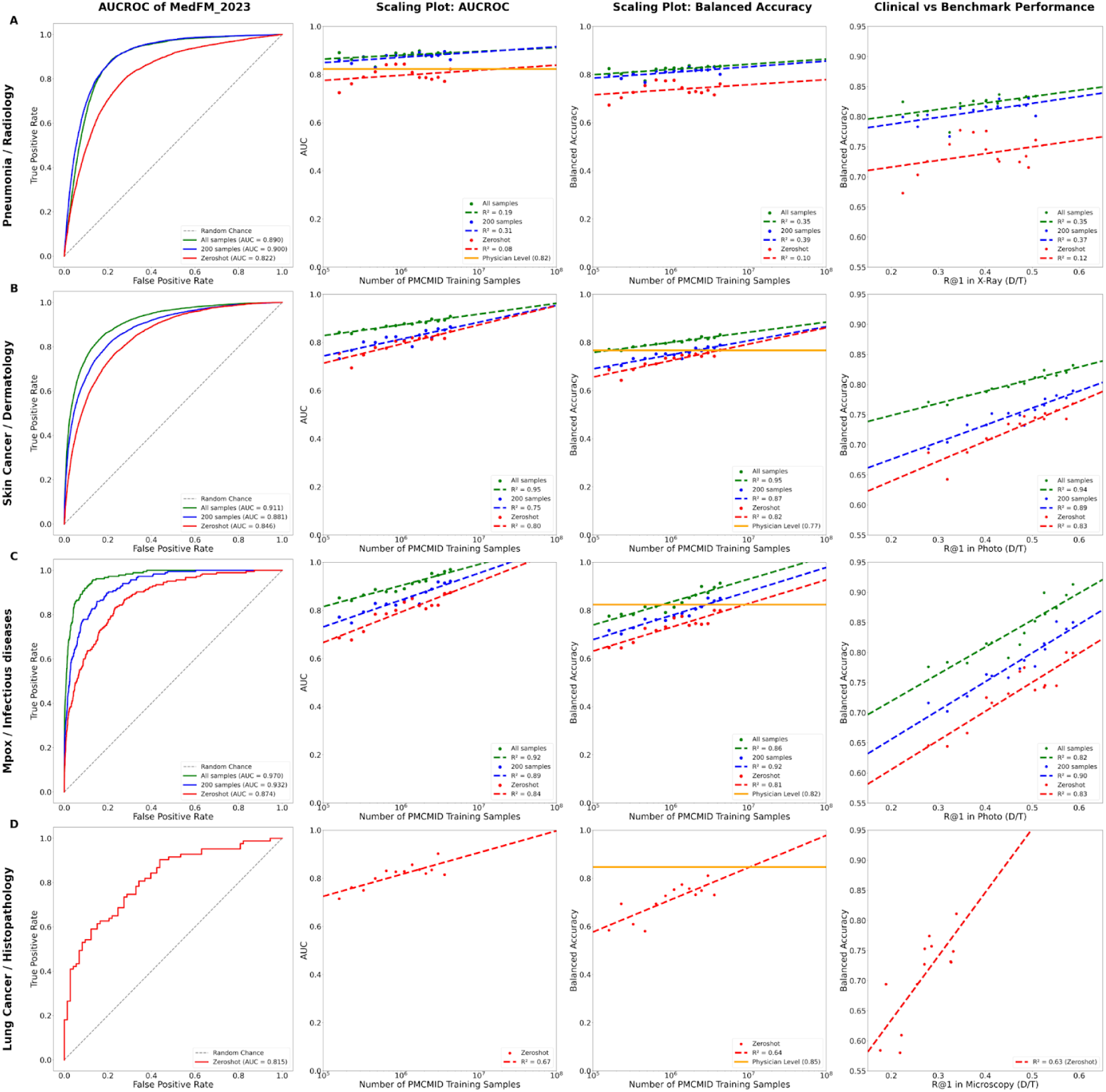
Performance of the MedFM in four different clinical classification tasks. A) Pneumonia detection in CRXs: MedFM_2023’s zero-shot performance (AUC=0.822) matched physician-level performance (AUC=0.823, 95% CI: 0.779-0.856). Scaling plots showed low to moderate correlations (Range R²: 0.12-0.39) between the number of PMCMID training images and clinical performance. There was a low to moderate correlation (Range R²: 0.12-0.35) between the performance in the PMCMID Benchmark test set in X-Ray images for diagnoses (R@1 in X-Ray (D/T) and the clinical performance. B) skin cancer detection in clinical photos: MedFM’s zero-shot performance (Balanced Accuracy: 0.768) matched physician-level performance (Balanced Accuracy 0.77). Scaling plots revealed a strong correlation (Range R²: 0.75-0.95) between the number of PMCMID training images and clinical performance (AUC and Balanced Accuracy), meaning that an increase in data in the PMCMID will likely result in a better clinical performance in this task. There was a high correlation (Range R²: 0.83-0.94) between the PMCMID Benchmark test set in Photos for diagnoses (R@1 in Photos (D/T)) and the clinical performance, meaning that an increase in performance as measured by the PMCMID Benchmark in Photos will likely result in an increase in clinical performance in skin cancer detection. C) mpox skin lesion detection in clinical photos: MedFM’s zero-shot performance (Balanced Accuracy: 0.799) matched physician-level performance (Balanced Accuracy: 0.82, 95% CI: 0.79-0.85). There was a high correlation in the scaling plots between the clinical performance and amount of training images in the PMCMID (Range R²: 0.81-0.92) and the clinical performance and the PMCMID Benchmark test set in Photos for diagnoses (Range R²: 0.82-0.90). D) Lung cancer subtyping in histopathology whole-slide images: MedFM_2022’s zero-shot performance (Balanced Accuracy: 0.811) approached the physician-level performance at 0.847. The scaling plot for balanced accuracy suggested (R²=0.64) that the MedFM could achieve physician-level performance with a PMCMID size of approximately 10.5 million training images. There was a moderate correlation (Range R²: 0.63) between the PMCMID Benchmark test set in Microscopy for diagnoses (R@1 in Microscopy (D/T)) and clinical performance.

We observed performance improvements in both zero-shot and CMLM-enhanced models as the number of PMCMID training images increased (**Fig. 5**, Scaling Plot: AUCROC, Scaling Plot: Balanced Accuracy). The AUCROC scaling plot showed a moderate R² of 0.31 with 200 domain-specific images. Similarly, we found moderate correlations of R² values of 0.35 and 0.39 with 200 and all available domain-specific images, respectively, in the balanced accuracy scaling plot (**Fig. 5A**, Scaling plot: AUCROC and Scaling plot: Balanced Accuracy). We observed a moderate correlation between balanced accuracy and R@1 in X-ray images of the PMCMID Benchmark with R² values of 0.37 and 0.35 for 200 and all samples, respectively (**Fig. 5A**, Clinical vs. Benchmark Performance).

### Detection of skin cancer in clinical photos

To assess MedFM’s ability to differentiate skin cancer - the most prevalent type of cancer worldwide^46^ - from other dermatological conditions in clinical photos, a dataset of 138,522 images from five public dermatological repositories (Danderm, DermIS, Hellenic Dermatological Atlas, DermNet, DermNet NZ), two public datasets (PAD-UFES-20^48^, Fitzpatrick 17k^49^), and a previously reported dataset (Esteva^50^**, Supplementary Table 7**) was compiled. We compared MedFM’s zero-shot capability with 38 medical terms (**Supplementary Table 8**) with a refined MedFM incorporating domain-specific images (Esteva cohort^50^), and a CMLM (**Fig. 1B**). MedFM’s zero-shot sensitivity and specificity of 0.769, and 0.768 matched physician-level performance of 0.798 (95% CI 0.732-0.851), and 0.736 (95% CI 0.665-0.796)^51^ (**Extended Fig. 1B** and **C**). Integrating MedFM_2023 with a CMLM improved the zero-shot performance of AUC=0.848 to 0.881 and 0.911 with 200 and 75,127 domain-specific images (**Fig. 5B, Supplementary Table 10, Supplementary Table 11**).

Balanced accuracy was 0.765 (95% CI: 0.759–0.771), increasing to 0.793 (95% CI: 0.789–0.797) with 200 domain-specific images and reaching 0.836 (95% CI: 0.836–0.836) using 75,127 domain-specific images. Scaling analyses showed strong correlations between model performance and the number of PMCMID training images, with R² values of 0.80–0.82 for zero-shot and 0.95 for CMLM-combined models. Balanced accuracy for skin cancer detection correlated highly with R@1 in photos of the PMCMID benchmark, with R² values of 0.83 for zero-shot and 0.94 for CMLM-combined models.

### Detection of mpox skin lesions in clinical photos

To evaluate MedFM’s ability to distinguish monkeypox virus (MPXV)-skin lesions from non-MPXV lesions, we used our previously published dataset of 139,198 photographic images (**Supplementary Table 12**)^54^. Performance was assessed in zero-shot mode and with a CMLM trained on domain-specific images from DermNetNZ, using 50 medical terms for mpox lesion classification (**Supplementary Table 13**).

MedFM_2023 achieved an AUCROC of 0.874 in zero-shot mode (**Supplementary Table 14**), increasing to 0.932 with 200 (**Supplementary Table 15**) samples and reaching 0.970 with all available domain-specific images (n=12,563) (**Supplementary Table 16**) (**Fig. 5C**). The balanced accuracy was 0.799 in zero-shot matching physician-level performance at 0.82 (95% CI 0.79-0.85). When deploying a CMLM, a balanced accuracy of 0.795 (95% CI 0.788-0.801) and 0.817 (95% CI 0.812-0.822) with 50 and 100 domain-specific images, respectively, could be reached (**Fig. 6C**).

With 200 samples, balanced accuracy reached 0.848 (95% CI: 0.843–0.852), surpassing physician-level performance at 0.913 when using all 12,563 domain-specific images (**Extended Fig. 1C**). Scaling analysis showed strong correlations between PMCMID training size and performance, with R² values of 0.84 (zero-shot), 0.89 (200 images), and 0.92 (all images) for AUCROC, and 0.81, 0.92, and 0.86 for balanced accuracy. MedFM_t (2010–2023) balanced accuracy correlated with PMCMID benchmark R@1 in photos, with R² values of 0.83 (zero-shot), 0.90 (200 images), and 0.82 (all images).

### Cancer subtyping of lung cancer in histopathological slides

Lung cancer is the leading cause of cancer-related deaths globally. Differentiating histological subtypes, lung adenocarcinoma (LUAD)^56^ and lung squamous cell carcinoma (LUSC)^57^, is critical for therapy. MedFM’s zero-shot capability in distinguishing LUAD (n=525) and LUSC (n=509) histopathology whole-slide images (WSIs) was evaluated using patch-based classification (**Fig. 1C**). Analysis revealed a progressive improvement in AUROC scores over time (PMCMID_t datasets 2011–2023, **Fig. 5D, scaling plot: AUCROC**). Starting from an AUROC of 0.715 (PMCMID_2011) to the model’s performance peak at 0.902 (PMCMID_2022, **Supplementary Table 17**). The performance of MedFM_2022 (Balanced Accuracy: 0.811) approached physician-level performance of 0.847^58^. Our projection using linear regression (R²=0.64) suggested that the MedFM could achieve physician-level performance with a PMCMID size of approximately 10.5 million training images (**Fig. 5D, Scaling plot: Balanced Accuracy**). We observed a moderate correlation between clinical performance and the PMCMID benchmark for microscopy images, with balanced accuracy for LUAD vs. LUSC classification showing a linear relationship with R@1 (R²=0.63). A CMLM was not used due to the lack of per-patch ground truth.

### Additional scaling plots

For all four clinical use cases, we created additional scaling plots for Accuracy, Error Rate, F1 Score, Precision, Sensitivity, and Specificity (**Supplementary Fig. 8-11**).

### Image2Paper App

The Image2Paper prototype app helps healthcare professionals interpret challenging visual findings by allowing users to upload or capture medical images via a smartphone (**Extended Fig. 3)**. It retrieves similar cases from 4.5 million PMCMID images (Type I–III) along with publication details, including the paper title, authors, figure caption, LLM-extracted diagnoses, and a PMC full-text link (**Extended Fig. 3C, 3D**). Similarity is determined using MedFM image embeddings, with results ranked by LLM-extracted diagnoses. In a clinical evaluation with five board-certified physicians, 43 images were tested. The correct diagnosis appeared in the top 10 results for 88.4% of cases and in the top three for 58.1%. Additionally, 93.0% of cases benefited from retrieved images and publications, providing valuable insights for differential diagnosis (**Supplementary Table 18).**

## DISCUSSION

We developed MedFM, a foundational vision-language model trained on the largest open-source annotated medical image dataset to date. MedFM achieved physician-level performance in classifying images across multiple medical specialties and modalities - without requiring additional training data. The model demonstrated an ability to comprehend relationships between imaging modalities, anatomical regions, and medical specialties. To assess robustness and potential biases, we introduced a manually annotated benchmark evaluating MedFM’s overall performance and its effectiveness across patient sub-cohorts. We identified scaling effects, showing that MedFM’s performance improves predictably with increasing training data. The Image2Paper app further illustrated MedFM’s versatility by reliably retrieving images with matching or related diagnoses, emphasizing potential real-world applications of MedFM.

The MedFM exhibits a general understanding of medical images, as evidenced in the following: UMAP analyses demonstrated MedFM’s ability to cluster medical images by modality (e.g., CT, X-rays), body region (e.g., abdomen, extremities), and specialty (e.g., cardiology, dermatology), highlighting its capacity to discern relationships across categories.

Image-text explanations using SHAP revealed that the MedFM consistently focused on image regions corresponding to pathological findings when making predictions toward the correct diagnosis across multiple imaging modalities and medical domains. In addition, when prompted with the term “normal,” the model identified the same regions but marked them as detracting from the prediction, indicating an understanding of the absence of pathology across diverse imaging domains. This dual capability suggested a new paradigm for medical classification: identifying deviations from normalcy rather than solely relying on condition-specific patterns. By focusing on identifying deviations from normalcy rather than condition-specific patterns alone, this approach could facilitate early detection of novel or rare conditions, where diagnostic labels might not yet exist, or to develop a general pathology/healthy classifier, capable of assessing whether a pathology is present, agnostic of the underlying condition.

MedFM achieved physician-level performance in three out of four cases in zero-shot mode and one approached physician-level with additional CMLM training. This highlighted the broad knowledge of the MedFM and its ability to generalize with high accuracy to clinical tasks and unseen data.

The Image2Paper app showcased the potential of the MedFM to apply its broad medical knowledge to an open-ended question by enabling clinicians to upload a medical image and retrieve similar images and publications from the Open Access medical literature. In our preliminary clinical evaluation, the tool offered a high accuracy in finding similar images with matching diagnoses or differential diagnoses. If validated in larger trials, Image2Paper could significantly impact medical practice, as it could assist physicians in validating their clinical assessments, particularly during tumor boards or other clinical meetings, by supplying tailored medical literature to the case being presented in real-time. Additionally, in cases where swift assessments are crucial and human expertise is scarce, Image2Paper could serve as an invaluable second opinion. Ultimately, this tool has the potential to enhance diagnostic accuracy and expedite both diagnosis and treatment in appropriate clinical settings and could provide a valuable resource for healthcare professionals.

We observed consistent scaling effects in MedFM’s performance across the PMCMID Benchmark and clinical tasks, with improvements correlating predictably with increased training data. In the PMCMID Benchmark, performance showed a strong correlation with the logarithm of training data size. Moderate scaling effects were noted in pneumonia detection and lung cancer subtyping, while skin cancer and mpox lesion detection exhibited strong correlations. These findings indicate that increasing training data size significantly enhances MedFM’s performance, with no evidence of a training plateau yet being reached.

With the exponential growth of medical images in PMCMID, automating its creation is essential to ensure regular updates. However, figure captions in medical publications often lack critical details for medical decision-making, such as diagnoses. We encourage future publications to include detailed, machine-readable descriptions, allowing each image and its peer-reviewed context to contribute to a comprehensive, annotated medical image database.

Additionally, incorporating further publicly available medical image datasets, such as The Cancer Imaging Archive (TCIA)^59^, MedPix^60^, or MURA^61^, would be desirable to enhance the number of training images significantly. Datasets like the UK-Biobank^62^ which primarily contain images of healthy individuals, could further enhance the model’s ability to differentiate between normalcy and pathological findings. Similarly, creating detailed image descriptions including diagnoses for each image in the datasets, as done with PMCMID, will be crucial to optimize the extraction of visual language knowledge from these sources.

A significant amount of public medical image-text pairs was available on the internet, yet not organized into structured datasets. Examples included the medical subset of Wikipedia^63^, Radiopaedia^64^, or WebPathology^65^. We hypothesized that MedFM could identify and filter relevant medical image-text pairs, similar to the approach used with the Large-scale Artificial Intelligence Open Network (LAION)^66^ dataset. This dataset, encompassing approximately 5 billion image-text pairs, was created by filtering web content using OpenAI’s CLIP model. By applying the same methodology, we proposed using MedFM to filter web content for medical image-text pairs, thereby compiling an even more extensive dataset of publicly available medical images, possibly one order of magnitude larger than the PMCMID. We anticipated retraining MedFM on this expanded dataset would significantly enhance its performance. Further, the volume of non-public medical data, such as that generated by hospitals, continued to grow steadily, offering a valuable resource of training data^67^.

A medical foundation model should be unbiased, ensuring fair outcomes across diverse populations. However, PMCMID reflected the imbalances present in the medical literature from which it was derived and, consequently, inherited these biases to the MedFM. Data biases in the medical literature are well known and may present important implications for social equity in healthcare^68^. Underrepresentation of groups such as patients with darker skin tones, pediatric populations, and those from resource-limited settings can result in performance inequities. For example, our Benchmark revealed a lower accuracy of the MedFM in patients with Fitzpatrick skin tones V-VI, underscoring the risk of exacerbating existing healthcare disparities. Scaling plots from the PMCMID Benchmark revealed that performance improvements for this patient group progressed significantly more slowly, with projections indicating that the disparities are likely to grow over time (**Extended Fig. 2D**).

Mitigating bias requires urgent action through several key strategies. Prioritizing the inclusion of vulnerable patient groups in research, data collection, and publications ensures better representation in medical datasets. Researchers should adopt inclusive study designs and transparently report demographic data to reduce biases in newly published data. Additionally, diversifying training datasets, integrating bias detection and correction mechanisms, and ensuring equitable representation in benchmarks are essential. These efforts aim to enhance the fairness, inclusivity, and reliability of medical foundation models, improving their applicability across diverse populations and clinical settings.

### Limitations

While MedFM demonstrated strong performance across four clinical use cases and in our multimodal benchmark, its generalizability across all image modalities and specialties remains uncertain. Computational limitations restricted full exploration of hyperparameter combinations despite optimization efforts. Our benchmark aimed to identify subgroup-specific biases, but some vulnerable populations, such as certain racial or ethnic minorities, may not be fully represented. Additionally, validation and test sets were annotated by 17 physicians, yet each image-text pair was reviewed by only one annotator due to dataset size. The Image2Paper app was tested with high-quality clinical images, not smartphone-captured images common in real-world settings, leaving its effectiveness in diverse clinical environments with variable image quality unverified.

## CONCLUSION

We developed MedFM, a generalist medical foundation model trained on PMCMID, the largest open-source annotated medical image dataset, covering all medical disciplines and imaging modalities. MedFM achieved physician-level performance across diverse clinical tasks without task-specific training, relying only on predefined medical terms. Performance improved with increasing training data, demonstrating strong scaling effects. The versatility of MedFM was further demonstrated through the Image2Paper app, enabling clinicians to retrieve similar images and relevant literature, supporting evidence-based decision-making. However, MedFM inherited biases from its training data, leading to disparities, particularly for underrepresented groups, such as patients with darker skin tones (Fitzpatrick V-VI). Projections indicated these disparities could widen over time. Therefore, we advocate for greater inclusion of vulnerable populations in research, transparent demographic reporting, and dataset diversification. We also emphasized the importance of including comprehensive AI-readable image descriptions in publications to support the ongoing development of medical foundation models. MedFM and PMCMID were made publicly available, with regular updates planned to ensure their sustained relevance and long-term impact on medical research and healthcare innovation.

## METHODS

### Ethics statement

Ethical oversight was provided by the Stanford institutional review board (Protocol: 36050, 67068 and 66980).

### Data sources

The PubMed Central Medical Image Dataset (PMCMID) comprises Open Access publications from PubMed Central (PMC; https://www.ncbi.nlm.nih.gov/pmc/) spanning from January 1, 2000, to January 30, 2023. A total of 16,126,659 images and their corresponding text (figure caption, referencing sentences, referencing paragraphs, title of the article, abstract, journal, and author information) were downloaded from PMC.

### Image selection and annotation

Given the extensive size of the PMCMID database, it was necessary to develop automated algorithms for image selection and annotation. We trained and used a convolutional neural network (CNN) to classify four types of images: Type I included medical images along with their descriptions; Type II featured panels of medical images and their descriptions; Type III combined panels of Type I and Type IV images; and Type IV non-medical images, e.g., abstract images, graphs, schematics. We created automatic annotation algorithms to enrich the figure caption with missing information to create comprehensive image descriptions. For this, we fine-tuned the Stanford BioMedLM^38^, a 2.7B parameter large language model trained on Pubmed abstracts and full articles, to identify and extract the diagnoses from the associated texts (title of publication and referencing paragraph to the image). We utilized a Unified Medical Language System (UMLS)^35^ parser to extract medical concepts from the associated texts. We extracted medical concepts related to 13 Unified Medical Language System (UMLS)^35^ semantic types: T019 for Anatomical Structure, T023 for Body Part, Organ, or Organ Component, T060 for Diagnostic Procedure, T047 for Disease or Syndrome, T033 for Finding, T058 for Health Care Activity, T037 for Injury or Poisoning, T170 for Intellectual Product, T048 for Mental or Behavioral Dysfunction, T191 for Neoplastic Process, T046 for Pathologic Function, T184 for Sign or Symptom, and T061 for Therapeutic or Preventive Procedure. We derived this list from a work that had previously identified UMLS semantic types related to medical decision-making^30,69^ and modified it for our task of visual knowledge extraction: 1) we replaced T029 (Body Location or Region) with T019 (Anatomical Structure) to enhance the specificity of the data annotations, 2) we removed T074 (Medical Device) as the imaging device was almost always mentioned in the figure caption, 3) we removed T200 (Clinical Drug) and T121 (Pharmacologic Substance), because drugs and pharmacological substances were mostly mentioned in the figure caption, if relevant, and are typically inferred from patient history rather from an image. We aimed to exclude images related to veterinary and animal research by excluding any publication where the journal title contained the term “veterinary” and any articles where the title referenced non-human subjects, specifically, “rat,” “dog,” “cat,” “mice,” “rabbit,” “cow,” “canine,” and “animal model”. We randomly selected 2% of Type I images and manually annotated them through a panel of multidisciplinary physicians (n=17) according to standardized annotation criteria **(Supplementary Methods, Extended Data Table 2**).

### Data splitting

Type I, II and III images were used for training, validation, or testing cohorts, and Type IV images were excluded. The manually annotated Type I images were used to create the PMCMID validation and test sets, each containing 1% of Type I images. All Type II and III images, and the remaining 98% of Type I images were used for the training cohort. To avoid data leakage across the training, validation, and test sets, we split the datasets by publication to ensure that images from the same patient, potentially appearing in multiple images of the same publication, did not cross over between sets. We created 14 subsets of the PMCMID dataset containing only data from publications till January 1 of each year from 2010 to 2023. These subsets are named PMCMID_t, where “t” denotes the year up to which data is included.

### Image processing and training algorithm

We utilized the OpenCLIP architecture, which combined a vision transformer (ViT) for image analysis and a transformer-based language model for text processing. Our approach involved systematically testing various training parameters to optimize model performance. Key parameters included:

- **Learning Rates:** {10^-4, 10^-5, 10^-6} for pretrained weights, {10^-3,10^-4} for training from scratch
- **Batch Sizes:** {128, 256, 512}
- **Number of Epochs:** {20, 30, 40, 50}
- **Image Types:** {Type 1, Types 1 & 2, Types 1, 2 & 3}
- **Vision Encoders:** {ViT-Base (both from scratch and pre-trained on LAION), ViT-Large (both from scratch and pre-trained on LAION), ViT-Huge (pre-trained on LAION)}
- **Text Augmentations:** {None, LLM diagnoses, LLM diagnoses + UMLS concepts from title, LLM diagnoses + UMLS concepts from title + referring paragraphs, LLM diagnoses + UMLS concepts from title + referring sentences}
- **Text Encoders:** {GPT-2 from scratch, PubmedBert, Stanford BioMedLM}

Due to the extensive training time of the MedFM for each parameter set, it was impractical to evaluate all possible combinations. We adopted a staged testing methodology, starting with a preliminary model configuration that exhibited promising performance. We then iteratively modified one parameter at a time to further enhance the model’s performance. This structured approach enabled us to effectively identify and refine the best-performing model configurations. We sourced the pre-trained weights for the image and text encoders from hugging face.

We trained the MedFM on an OpenCLIP architecture on supercomputing resources provided by JUWELS^37^. We found the following training parameters to result in the highest R@1 in all images of the validation cohort: The visual encoder backbone was VIT-Large pre-trained on LAION all layers unlocked for training, the text encoder backbone PubMedBert pre-trained on Pubmed with four unlocked layers, training dataset consisted of all image types containing medical images (Type 1,2, and 3), data augmentation using the TIMM library with the parameters “’scale’: (0.3, 1.0), ‘re_prob’: 0.4”, lr = 10-e5, epochs = 40, warmup = 2000, batchsize = 256 (in a 16 GPU configuration), wd = 0.2, eps 1e-6, local-loss.

After identifying the best training parameters, we trained the Medical Foundation Model at time point t (MedFM_t) on the image-text pairs within each of the 14 PMCMID_t datasets. This allowed us to assess the impact of increasing data availability on model performance.

### Algorithm evaluation

#### PMCMID Benchmark algorithm

The objective of the PMCMID Benchmark was 1) to guide us during the training process of the MedFM to identify optimal training parameters and 2) assess the performance of MedFM across various sub-cohorts, including image modalities, age groups, sexes, body regions, and skin tones.

The Benchmark algorithm is based on the ability of the MedFM to calculate cosine similarities between medical images and text, which enabled the algorithm to rank multiple texts according to their cosine similarity values. This enabled measuring the performance as R@N (Recall at Rank N), which is the ratio of images whose correct image description was ranked within the top N ranks. R@1 measures the proportion of images for which the correct image description is ranked as the top result (rank 1) based on cosine similarity, while R@10 indicates the fraction of images with the correct description appearing within the top ten ranks. In general, the following formula is used to calculate R@N: R@N = number of correct image-text matches at ranks ≤ N / total number of image-text pairs.

With an increasing total number of image-text pairs, this image-to-text matching task becomes increasingly more difficult for the model. Varying sizes of subgroups (e.g., gender, skin type, body regions, image modalities, and age) would introduce inconsistencies between the performance measurements and would not allow to compare performances between groups. For this reason, we standardized the image-to-text matching task to contain 50 random image-text pairs and repeated the process 100 times to calculate averages for R@1 and 95% CI intervals. For each iteration, ranks for each image were calculated, and the ranks were then aggregated across all iterations. Confidence intervals for R@N metrics were calculated using the Bootstrapping method.

We analyzed R@1 with 95% CIs, to calculate p-values for the differences in group performances from a Bonferroni-corrected Mann-Whitney U test (**Fig. 2A-B**). Finally, to create benchmarks of higher difficulty and at the expense of losing subgroups due to their insufficient size, R@1 was evaluated against a higher number of P (with P being the total number of image-text pairs) by incrementally increasing P from 50 to the maximum for each group. All analyses were conducted within the image-to-text R@N context, with the text comprising either full image descriptions (including diagnoses) or diagnosis-only descriptions.

### Zero-shot

MedFM was capable of zero-shot learning to recognize and classify medical conditions that it had not been explicitly trained to identify. This was achieved by leveraging a comprehensive medical knowledge base acquired during training, which the model applied to new, unseen categories of data. Our zero-shot classification process started by gathering a list of relevant medical terms, including the names of the disease to be classified and its variants, visual symptoms, the most common differential diagnoses (in cases where diseases were mutually exclusive), the anatomical region(s) when the disease had a predilection for certain areas, and descriptions of what constitutes normality (absence of the disease). These terms were sourced from a panel of physicians. For classification purposes, these terms were then organized into ‘positive’ and ‘negative’ categories. ‘Positive’ terms directly related to the presence of the specific medical condition, while ‘negative’ terms related to its absence or to distinguishing features of alternative diagnoses. For each image-text pair, MedFM calculated the cosine similarities between the image features and the embeddings of each medical term, resulting in a set of logits. Given that our list of medical terms included disease variants and visual symptoms - which may co-occur rather than being mutually exclusive - the traditional softmax function, which assumed exclusivity and normalized probabilities across all classes to sum to one, was not suitable. Instead, we used a modified softmax function. We calculated the average of the logits belonging to the same medical condition within the positive and negative categories, creating aggregated scores for each medical condition. We chose the medical conditions in such a way, so they were mutually exclusive. We then applied the softmax function to the aggregated scores, transforming them into probabilities for each medical condition. This methodology allowed to query MedFM on multiple medical terms and harness its broad medical knowledge in a zero-shot manner, effectively generalizing to unseen image-text pairs without relying on direct, example-based training.

Mathematical model of the modified softmax function:

1. MedFM calculated the cosine similarities between each image i and each medical term *j*, creating logits *l_ij:_*

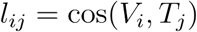

1. Aggregation of Logits: For each medical condition within the positive and negative categories:

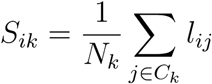

Here, *S_ik_* represented the aggregated score for condition *k* for image-text pair *i*, where *N_k_* was the number of terms *j* associated with condition *k* in category *C_k_* (either positive or negative).

1. Modified Softmax function:

The modified softmax function was applied to convert the aggregated scores *S_ik_* into probabilities:

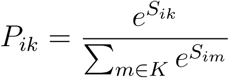

Where *P_ik_* was the probability that the image-text pair *i* belonged to medical condition *k*. The sum in the denominator ran over all medical conditions *m* being considered.

### Refinement of MedFM’s performance using domain-specific images and Classical Machine Learning Models (CMLMs)

The modified softmax function used by MedFM operated under several key assumptions:

1. **Single-Condition Association for Medical Terms:** When aggregating the scores of each medical term, the function assumed that each term is associated with only one medical condition. However, many medical terms, such as symptoms, are not pathognomonic and may be linked to multiple conditions.
2. **Equal Weighting of Logits:** By averaging logits, the function assumed that all medical terms have equal influence on category predictions. This approach overlooked the possibility that certain terms might carry greater diagnostic weight or be more critical for identifying specific conditions.
3. **No Interactions Between Terms:** We presumed that medical terms and their corresponding logits functioned independently, ignoring potential interactions. This simplification failed to account for the synergistic or antagonistic effects often observed when combinations of symptoms or findings are considered in medical diagnostics.

Given these limitations, we explored the potential benefits of replacing the modified softmax function with a classical machine learning model (CMLM). By training the CMLM on a dataset of domain-specific images with known ground truth separate from PMCMID, we aimed to capture potential improvements in MedFM’s performance using this technique. For the selection of the optimal machine learning model, we employed a cross-validation and hyperparameter tuning approach using RandomizedSearchCV from Scikit-learn. Our model suite included Logistic Regression, Random Forest, Gradient Boosting, and Support Vector Machines (SVM), each evaluated across a spectrum of hyperparameter settings to find the most effective configuration. The hyperparameters for Logistic Regression were tuned over a range of regularization strengths with C values from np.logspace(−2, 2, 10). For Random Forest, we explored hyperparameters including n_estimators set to 1000, max_depth options of None, 10, 20, and 30, min_samples_split choices of 2, 5, and 10, and class_weight settings of ‘balanced’, ‘balanced_subsample’, and None.

Gradient Boosting hyperparameters included n_estimators of 50, 100, and 150, learning_rate values of 0.01, 0.1, and 0.2, and max_depth of 3, 5, and 7. For SVM, the C parameter was sampled from a log uniform distribution over a range from 1e-3 to 1e3, kernels included ‘linear’, ‘poly’, ‘rbf’, and ‘sigmoid’, gamma values were drawn from a log uniform distribution spanning 1e-4 to 1e1, degrees were set to 2, 3, and 4, and coef0 was explored with uniform(0, 10).

The cosine similarities calculated by the MedFM from the input image and medical terms were fed into different CNMLs to train on predicting the target condition with a varying number of domain-specific images (50, 100, 200, full dataset). Hyperparameter optimization was conducted through K-Fold cross-validation with 5 splits, ensuring the reliability of our model assessments. We randomly sampled 100 different hyperparameter settings per model to evaluate their performance, focusing particularly on maximizing the AUCROC for diagnostic accuracy. The best performing model and its parameters were selected based on the highest AUCROC score obtained during the validation process.

### Image-Text Explainability

We employed Shapley Additive Explanations (SHAP) to elucidate image-text correlations. To assess which image regions were most informative of the text description, we used an iterative image masking approach. Specifically, we perturbed the input images by applying a 20×20 pixel Gaussian blur to mask regions, simulating feature removal while preserving the overall structure of the image. For each perturbation, we measured the resulting change in the model’s predicted probability for the corresponding text output, attributing the impact to individual pixel regions. To balance computational efficiency and granularity, on each image we set the maximum number of model evaluations to 5,000 during the SHAP value computation. Similarly, to assess which parts of the text description were most relevant to the image input, we used a text masker to iteratively mask out each vocabulary within the text input.

### UMAPs

To display and analyze the image and text embeddings, UMAPs^39^ were created. Each data point, initially represented as a high-dimensional vector embedding corresponding to either an image or an image description, was reduced to a two-dimensional point through an advanced dimensionality reduction technique called UMAP. This process preserves both global and local structures of the high-dimensional data within a simplified 2D space. The interactive UMAP visualizations (**Code availability**) allowed users to hover over each UMAP data point to reveal relevant information and metadata linked to the embeddings, such as the image itself, the text description, body region, medical image modality, and medical specialty.

### Detection of pneumonia in chest X-Ray (CRX) images

To assess the performance of the MedFM in distinguishing pneumonia from other conditions in CRX images, we curated a dataset from four large CRX collections: CheXpert^42^, MIMIC-CRX^43^, PadChest^44^, and RSNA^45^ (**Supplementary Table 2**). These datasets comprise 224,316; 377,110; 160,828; and 30,277 images, respectively, with most annotations generated automatically via NLP from radiology reports and a smaller subset manually verified by physicians.

We created a test dataset from physician-annotated CRX images in AP or PA projection across all datasets, identifying 5,885; 26,432; 2,004; and 30,227 images in CheXpert, MIMIC-CRX, PadChest, and RSNA, respectively, and a total of 64,548 CRX images.

We evaluated the zero-shot performance of MedFM. In addition, we aimed to increase the performance of the MedFM by replacing the softmax function with a CMLM (**Fig. 1B**, Whole-image classification). We evaluated the performance of the CMLM with a varying number of training images, which we randomly sampled from the CheXpert training cohort (NLP annotated). We randomly sampled for 20 iterations 50, 100, and 200 samples from the training cohort and calculated the average performance and 95% CIs. To get the best performance, we used the entire training cohort (all samples=5885) for training the CNML. The list of medical terms used for this evaluation can be found in **Supplementary Table 3.**

For scaling plots, we created several instances of the foundation model MedFM_t (with t ranging from 2010 to 2023), which were trained on corresponding PMCMID_t datasets. This setup allowed the analysis of how an increase in the number of PMCMID training images influenced model performance. We observed increasing performance in both zero-shot and with CMLMs as the number of PMCMID training images increased, reflected in both AUCROC and balanced accuracy metrics.

To research the question, whether the performance in the PMCMID benchmark correlated to the clinical performance in this task, we plotted the performance for all MedFM_t from 2010 to 2023 for the balanced accuracy in the CRX dataset vs. the R@1 in X-Ray in the PMCMID Benchmark.

We obtained the physician baseline AUROC of 0.823 (95% CIs: 0.779-0.856) from a study, which assessed the performance of three cardiothoracic radiology specialists on a 420-image sample from the NIH-CRX8 dataset^70^.

### Detection of skin cancer in clinical photos

To evaluate the MedFM’s ability to distinguish skin cancers from other dermatological conditions in clinical photos, we compiled a dataset of 138,522 images from five public dermatological repositories (Danderm, DermIS, Hellenic Dermatological Atlas, DermNet, DermNet NZ), two public datasets (PAD-UFES-20^48^, Fitzpatrick 17k^49^), and one institutional dataset (Esteva^50^). We used Danderm, DermIS, Hellenic Dermatological Atlas, DermNet, DermNet NZ, PAD-UFES-20, and Fitzpatrick 17k as testing cohorts and Esteva as training cohort. We evaluated the performance of the MedFM in zero-shot and in combination with a CMLM with varying numbers of samples from the training cohort 50, 100, 200, and 75,127 samples. The list of medical terms can be found in **Supplementary Table 8**. We created scaling plots for AUC and balanced accuracy analogously to the pneumonia cohort. To correlate the clinical performance vs. the benchmark performance, we compared balanced accuracy with R@1 in photo (D/T) of the benchmark cohort for all MedFM_t with t ranging from 2010 to 2023. For physician-level performance, we obtained a pooled sensitivity of 79.8% (95% CI 73.2-85.1%) and a pooled specificity 73.6% (95% CI 66.5-79.6%) for physicians from a meta-analysis^51^ and calculated a physician-level balanced accuracy of 76.7%.

### Detection of mpox skin lesions in clinical photos

To assess the performance of MedFM_2023 in distinguishing MPXV-skin lesions from non-MPXV lesions, we utilized a dataset of 139,198 photographic images, as detailed in^54^. For MPXV-skin lesions we sourced the images from publications of the medical literature, social media, encyclopedia, news articles, and a prospective cohort of patients treated at Stanford University. For non-MPXV lesions, we used Danderm, DermIS, Hellenic Dermatological Atlas, DermNet, Esteva, PAD-UFES-20, and Fitzpatrick 17k as testing cohorts and DermNet NZ as training cohort. We evaluated the performance of the MedFM in zero-shot and in combination with a CMLM with varying numbers of samples from the training cohort 50, 100, 200, and 12,563 samples. We created scaling plots for AUC and balanced accuracy and correlated the clinical performance with the benchmark performance analogously to the skin cancer cohort. For mpox detection in photographic images, we were unable to find a physician-level performance in the literature. For this reason, we shared a sample of 100 images of our dermatology database with 10 physicians who manually labeled the dataset. We experimentally obtained a physician-level performance of a balanced accuracy of 0.82 (95% CI 0.79-0.85).

### Cancer subtyping of lung cancer in histopathological slides

To evaluate the MedFM’s performance on histopathology images, whole-slide images (WSIs) of both Lung Adenocarcinoma (LUAD)^56^ and Lung Squamous Cell Carcinoma (LUSC)^57^ of the dataset provided by the National Cancer Institute (NCI) Genomic Data Commons (GDC)^71^ were tested. Following the methodology by Coudray et al.^58^, a balanced set of n=1034 ‘primary tumor’ slides (LUAD: n=525 WSIs; LUSC: n=509 WSIs) were extracted. We replicated their approach by splitting the dataset into training (70%), validation (15%), and testing (15%) sets using a random split method. Additionally, data preprocessing was performed. Precisely, 100 tiles (measuring 512 x 512 pixels) of WSIs were extracted at level 2 of the WSIs, corresponding to a native magnification factor of 2.5X. Each extracted tile measures 512 x 512 pixels. This tiling process was performed randomly across the slides to capture a diverse set of features inherent to the tissue samples. The randomly selected tile regions are configured to contain a minimum of 80% tissue coverage, validated through automated tissue detection methods, including grayscale conversion and Otsu thresholding^72^. 100 tiles were extracted from each WSI.

The predictive analysis was conducted on two levels: tile and patient. Initially, the MedFM was tasked with classifying each tile as LUAD, LUSC, or normal tissue. For this purpose, we prompted the MedFM to the tile-level predictions that were aggregated to generate patient-level outcomes, utilizing the best aggregation strategy determined through validation set performance. This approach aimed to reflect the MedFM’s ability to diagnose lung cancer subtypes, akin to clinical settings accurately.

At the patch level, we computed cosine similarities between the model’s output embeddings and embeddings of descriptive labels for each class, including LUAD, LUSC, and normal lung tissue. Using the scoring mechanism based on cosine similarities enabled the quantification of alignment between the image and each possible label. For aggregating tile-level predictions to a patient-level diagnosis, we first excluded scores for the normal tissue label and identified the tiles with the top 10 scores for LUAD or LUSC, and then calculated the average score across all labels to find the label with the highest score to be the patient-level prediction of the model.

To evaluate the predictive performance, we computed key metrics such as accuracy, F1 score, balanced accuracy, class-specific precision, recall, specificity, and their macro averages. Furthermore, we benchmarked the model’s performance against clinical standards by comparing it to the performance of three pathologists, as documented in the referenced study. Physician performance metrics were derived from the confusion matrix provided in that study^58^.

### Image2Paper

We computed embeddings for roughly 4.5 million images from the PMCMID dataset using the MedFM, which was trained on a diverse set of medical images accompanied by detailed descriptions. We hypothesized that this training approach, allowed the MedFM to discern subtle similarities and differences among images, focusing on underlying pathologies and medical concepts.

For each uploaded image, Image2Paper (**Extended Figure 3**) performed an image to image comparison using the MedFM by comparing the image embeddings of the input image with image embeddings of all images in the PMCMID. After the 30 most similar medical images are identified, the MedFM was used to calculate the cosine similarities between the input image and the LLM-extracted diagnoses of the identified images. Images were then ranked after the cosine similarity of their diagnosis and the input image. The top 10 of these images were then presented to the user. We integrated this functionality into a Django-based web application, enabling access via browsers on smartphones and allowing direct photo uploads through the smartphone’s camera. The web server with an Intel Xeon Platinum 8358 and a NVIDIA A100 GPU could complete a request in under a second.

We conducted a clinical evaluation with five physicians, asking each to upload between 5 to 10 images of known conditions to the app. After each upload, they completed a survey indicating whether the correct diagnosis appeared among the top 10 results, its specific ranking, and the presence of differential diagnoses. The physicians also provided feedback on the utility of the app outputs in their clinical practice and explained their reasons.

## DATA AVAILABILITY

### Data Availability for PMCMID

Dataset will be released prior to publication and will contain metadata for each medical image. Since we do not hold copyright for the images, we are unable to include the images directly into the dataset and make them available for download. However, the metadata contains the link to the publication archives and images files within the archives. We will provide a script for downloading images from PMC servers allowing researchers to generate their own PMCMID datasets. This enables researchers to independently compile the dataset for their specific needs.

## CODE AVAILABILITY

Code for training the MedFM and Benchmark algorithms will be provided prior to publication (GitHub Repository).

**Link to interactive UMAPs:** http://74.208.243.67/allUMAPs.html

## Author Contributions

A.H.T. designed the research, designed and developed the MedFM, collated and analyzed the data, constructed the PMCMID, designed, developed, and annotated the PMCMID benchmark dataset, designed and developed benchmark algorithms, designed and developed the Image2Paper algorithm and app and wrote the first draft of the manuscript. T.M. evaluated the MedFM, designed and developed benchmark algorithms, designed and developed interactive UMAPs, analyzed the data, created graphics, and wrote the manuscript. M.G., A.R.M., T.K., G.R., J.S., and J.B. annotated the PMCMID benchmark dataset, evaluated the Image2Paper app, provided important intellectual input, and reviewed the manuscript. Y.Z. designed and developed explainability and reviewed the manuscript. K.P. designed and developed the webapp interface of the Image2App app, analyzed the data, and reviewed the manuscript. G.M. created graphics and reviewed the manuscript. M.M. collated and analyzed the data, designed, developed, and annotated the PMCMID benchmark dataset, evaluated the Image2Paper app, created graphics, aided in the development of methods and interpretation of results, and wrote the manuscript. O.G. aided in the development of methods and interpretation of results and reviewed the manuscript.

## Data Availability

All data produced in the present study are available upon reasonable request to the authors.

## Acknowledgments

A.H.T. and M.M. are fellows of the BIH - Charité Digital Clinician Scientist Program funded by the Charité - Universitätsmedizin Berlin, the Berlin Institute of Health at Charité, and the German Research Foundation (DFG). A.R. was supported by a Clinician Scientist Program of the Medical Faculty of the University of Leipzig.

The authors gratefully acknowledge the Gauss Centre for Supercomputing e.V. (www.gauss-centre.eu) for funding this project by providing computing time on the GCS Supercomputer JUWELS at Jülich Supercomputing Centre (JSC).

## Extended Data

**Extended Figure 1:**
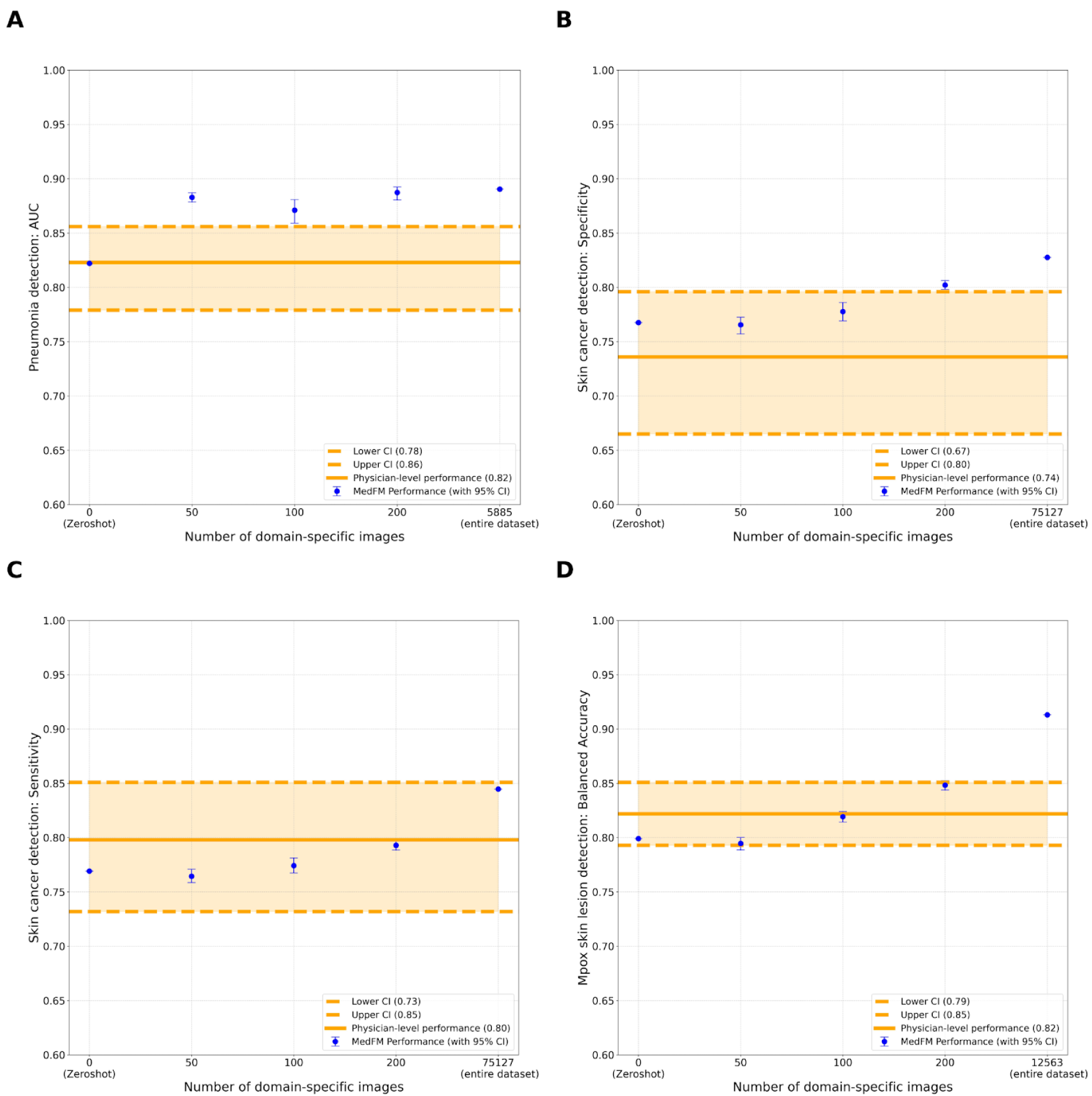
Comparison of MedFM’s Performance Against Physician-level Performance. Comparison of MedFM’s performance in zero-shot and when combined with a Classical Machine Learning Model (CMLM) trained on various numbers of domain-specific images, against physician-level performance. A) Pneumonia Detection in CRX: MedFM achieved a zero-shot AUC of 0.822, closely matching physician-level (AUC=0.823, 95% CI: 0.779-0.856). When we combined the MedFM with a CMLM that we trained on 50 domain-specific CRX images, MedFM’s performance increased to an AUC of 0.883 (95% CI: 0.879-0.887), surpassing physician-level performance. B and C) Skin cancer detection in clinical photos: In zero-shot, MedFM had a sensitivity and specificity 0.77 and 0.77 and matched physician-level performance of sensitivity 0.798 (95% CI 0.732–0.851), and specificity of 0.736 (95% CI 0.665-0.796). Above physician-level performance was achieved with a specificity of 0.828 when MedFM was combined with a CMLM trained on 75,127 domain-specific images C) Mpox detection in clinical photos: MedFM’s balanced accuracy was 0.799 in zero-shot matching physician-level performance (0.82, 95% CI: 0.79-0.85). Above physician-level performance was achieved with a balanced accuracy of 0.913 when MedFM was combined with a CMLM trained on 12,563 domain-specific images.

**Extended Figure 2:**
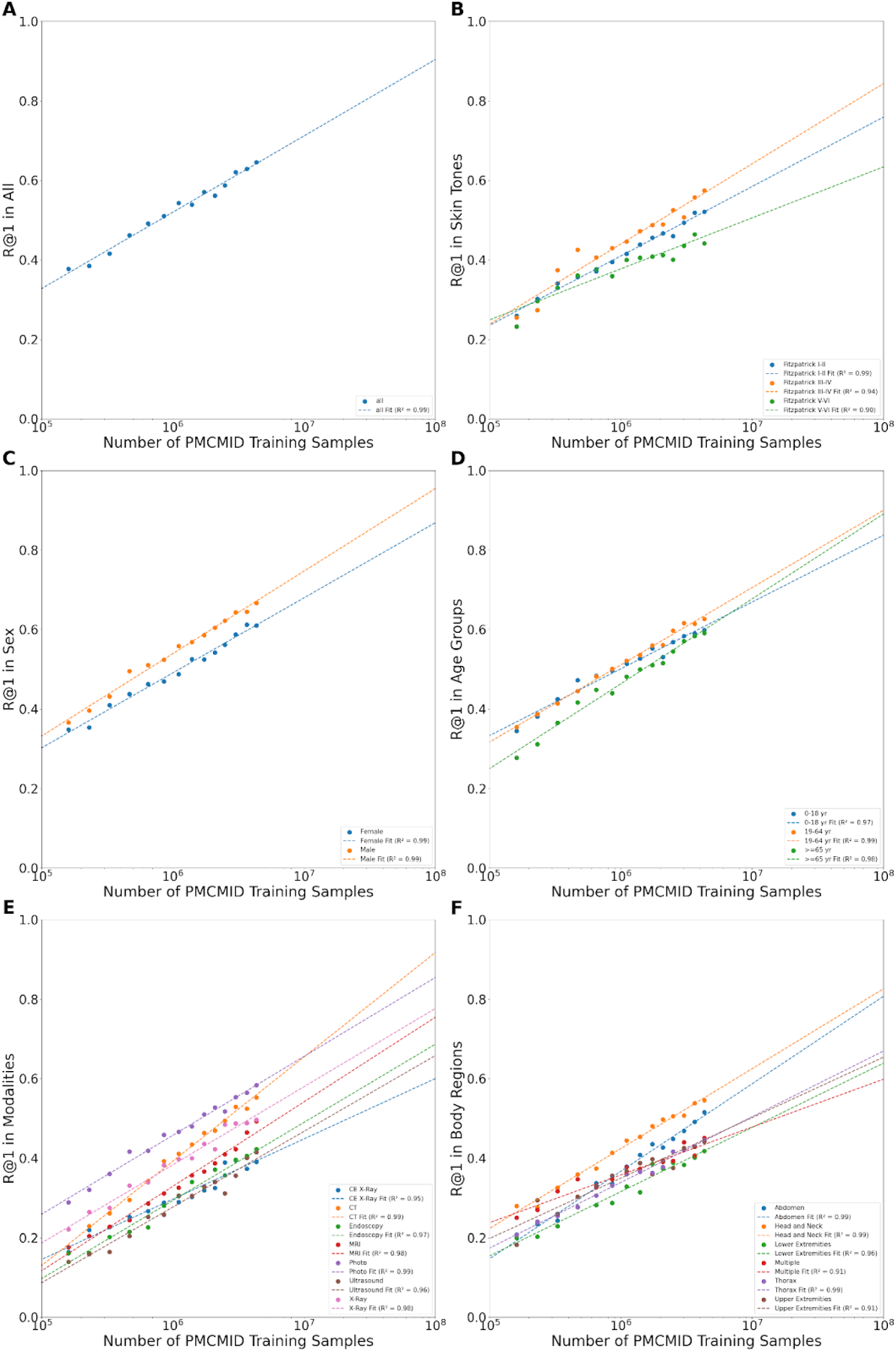
Scaling plots of MedFM’s performance in the PMCMID testing cohort. Each panel displays the relationship between performance, measured using the R@1 metric, and the number of PMCMID training samples on a logarithmic scale. A strong correlation (R² = 0.90-0.99) is observed across all panels, indicating that MedFM’s performance improvements can be accurately predicted as a function of training data size: A: R@1 for all images shows a strong correlation (R² = 0.99) with the number of training samples, underscoring predictable performance scaling with increased data. B: R@1 performance for skin tone groups (Fitzpatrick I-II, III-IV, V-VI). Fitzpatrick V-VI consistently underperforms relative to lighter skin tones. Projections suggested that performance gaps between skin tone groups could widen further as training data increased due to differences in improvement rates. C: R@1 performance by sex: Male patients consistently outperform female patients. However, projections indicated no significant differences in the slopes of performance improvement, suggesting that the existing performance gap is unlikely to worsen substantially with additional training data. D: Performance by age groups. Older age groups (≥65 years) demonstrated slower initial performance but improved at a higher rate than other age groups. E: Performance by imaging modality. Differences in performance across modalities were observed with projections suggesting that performance in CT images increased at a higher rate. F: Variability across body regions (e.g., abdomen, head and neck, thorax) highlighted differences in MedFM’s ability to generalize across anatomical locations.

**Extended Figure 3:**
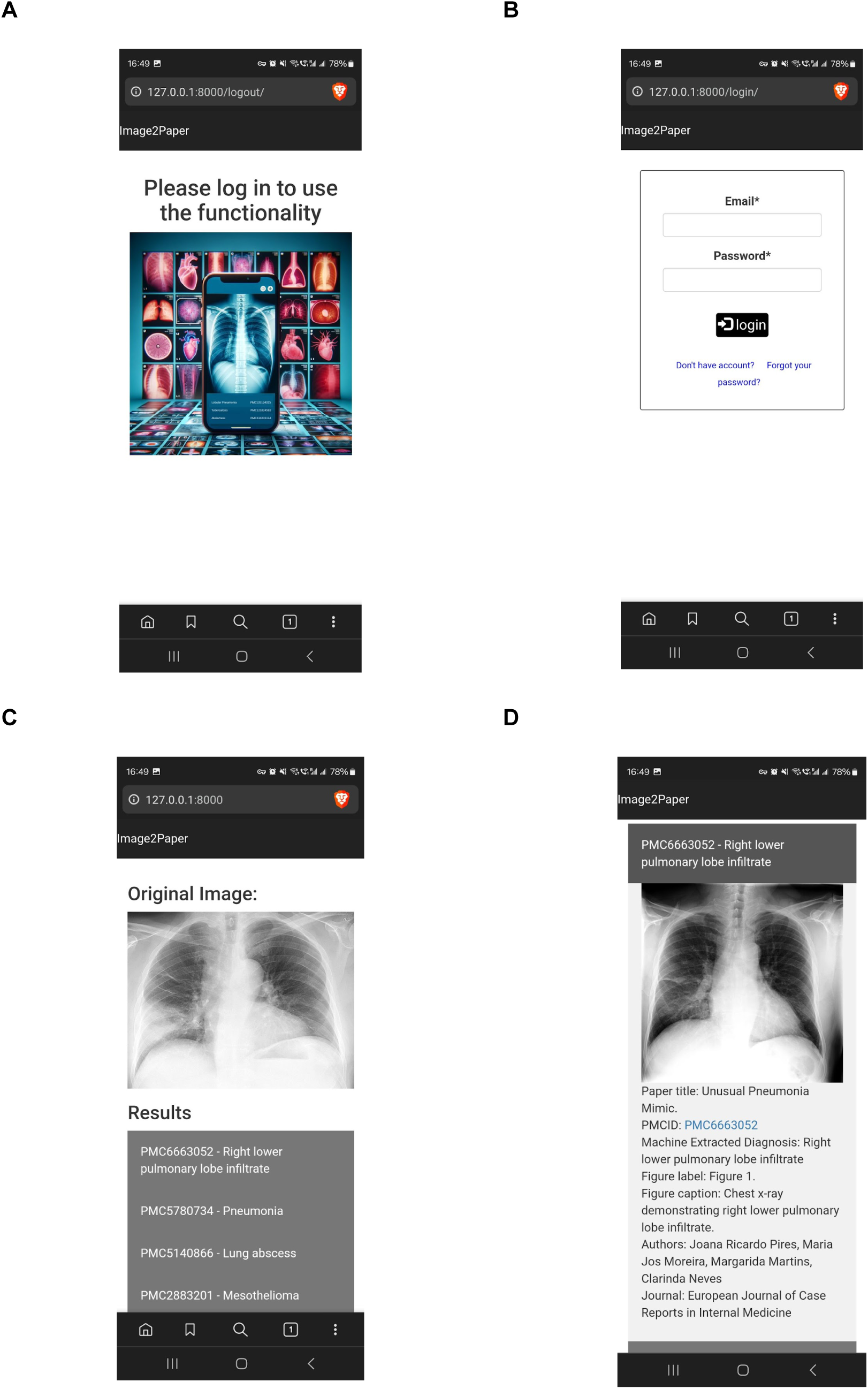
Screenshots of the Image2Paper App Prototype. A) Welcome page with initial user interface. B) Login page for user authentication. C) Uploaded image featuring a chest radiograph (CRX) of right lower pneumonia, along with associated publications identified by the app. D) Detailed view of the first identified publication, showing the publication image, publication title, LLM-extracted diagnosis, figure caption, journal, list of authors, and a PMC full-text link.

**Extended Data Table 1:** Number of open-access images available from PubMed publications per image type and year. Type I images are single medical images with one description; Type II images consists of a panel of medical images and their descriptions; Type III images are a panel of Type I and Type IV images and their descriptions; and Type IV images are abstract images, graphs, schematics etc.

| Dataset (PMCMID_YYYY) | End-Date | All Images (n) | Type 1 Images (n) | Type 2 Images (n) | Type 3 Images (n) | Type 4 Images (n) | Type 1,2, and 3 Images (n) | Images excluded for training (n) | PMCMID Training Images (n) | Model (MedFM_YYYY) |
| --- | --- | --- | --- | --- | --- | --- | --- | --- | --- | --- |
| - | 2000-01-01 | 27721 | 1157 | 5104 | 4584 | 16876 | 10845 | 17300 | 10410 | - |
| - | 2001-01-01 | 34362 | 1395 | 6437 | 5927 | 20603 | 13759 | 21086 | 13265 | - |
| - | 2002-01-01 | 41159 | 1648 | 7829 | 7451 | 24231 | 16928 | 24779 | 16369 | - |
| - | 2003-01-01 | 49793 | 2105 | 9257 | 9266 | 29165 | 20628 | 29776 | 20001 | - |
| - | 2004-01-01 | 59286 | 2586 | 10682 | 11401 | 34617 | 24669 | 35302 | 23968 | - |
| - | 2005-01-01 | 76487 | 3564 | 12833 | 14310 | 45780 | 30707 | 46628 | 29827 | - |
| - | 2006-01-01 | 107155 | 5331 | 16308 | 18829 | 66687 | 40468 | 67801 | 39299 | - |
| - | 2007-01-01 | 153646 | 8242 | 21119 | 25149 | 99136 | 54510 | 100772 | 52766 | - |
| - | 2008-01-01 | 224629 | 13091 | 28934 | 35447 | 147157 | 77472 | 149453 | 74972 | - |
| - | 2009-01-01 | 341648 | 21294 | 41578 | 51384 | 227392 | 114256 | 230705 | 110591 | - |
| 2010 | 2010-01-01 | 504362 | 33758 | 59090 | 73301 | 338213 | 166149 | 343033 | 160766 | 2010 |
| 2011 | 2011-01-01 | 733599 | 49697 | 84872 | 104827 | 494203 | 239396 | 501457 | 231351 | 2011 |
| 2012 | 2012-01-01 | 1047216 | 70639 | 122545 | 148437 | 705595 | 341621 | 716081 | 330034 | 2012 |
| 2013 | 2013-01-01 | 1491098 | 98616 | 175849 | 210780 | 1005853 | 485245 | 1021451 | 468100 | 2013 |
| 2014 | 2014-01-01 | 2073776 | 131325 | 247596 | 293740 | 1401115 | 672661 | 1423527 | 648173 | 2014 |
| 2015 | 2015-01-01 | 2775042 | 169448 | 332047 | 389147 | 1884400 | 890642 | 1914239 | 858263 | 2015 |
| 2016 | 2016-01-01 | 3602443 | 213679 | 429635 | 505571 | 2453558 | 1148885 | 2492540 | 1106765 | 2016 |
| 2017 | 2017-01-01 | 4581357 | 258315 | 541845 | 650586 | 3130611 | 1450746 | 3179622 | 1397964 | 2017 |
| 2018 | 2018-01-01 | 5725566 | 306575 | 671093 | 822124 | 3925774 | 1799792 | 3986567 | 1734575 | 2018 |
| 2019 | 2019-01-01 | 7038419 | 356634 | 815302 | 1001507 | 4864976 | 2173443 | 4937827 | 2095551 | 2019 |
| 2020 | 2020-01-01 | 8548439 | 409506 | 974112 | 1201732 | 5963089 | 2585350 | 6048875 | 2493887 | 2020 |
| 2021 | 2021-01-01 | 10554425 | 484287 | 1189568 | 1449975 | 7430595 | 3123830 | 7532812 | 3014957 | 2021 |
| 2022 | 2022-01-01 | 13160534 | 570851 | 1457519 | 1755245 | 9376919 | 3783615 | 9498187 | 3654626 | 2022 |
| 2023 | 2023-01-01 | 16104074 | 656297 | 1743443 | 2076923 | 11627411 | 4476663 | 11769003 | 4326353 | 2023 |
| - | 2023-01-30 | 16126659 | 656818 | 1745444 | 2080065 | 11644332 | 4482327 | 11786128 | 4331810 | - |

**Extended Data Table 2:** Characteristics of the validation and test set. This table describes the number of all images, and the number of sub groups (e.g. number of images with a specific skin tone) in the validation and test set.

|  | Validation | Test | Total |
| --- | --- | --- | --- |
| All | 6730 | 6536 | 13266 |
| Included | 4470 | 4268 | 8738 |
| Excluded | 2260 | 2268 | 4528 |
| Sex |  |  |  |
| Female | 857 | 920 | 1777 |
| Male | 930 | 1097 | 2027 |
| N/A | 57 | 42 | 99 |
| Unknown | 2626 | 2209 | 4835 |
| Age |  |  |  |
| Child (0-18 years) | 360 | 290 | 650 |
| Adult (19-64 years) | 883 | 895 | 1778 |
| Senior (>= 65 years) | 279 | 315 | 594 |
| Unknown | 2888 | 2727 | 5615 |
| N/A | 60 | 41 | 101 |
| Skin Tone (Fitzpatrick) |  |  |  |
| I | 60 | 78 | 138 |
| II | 263 | 227 | 490 |
| III | 155 | 114 | 269 |
| IV | 139 | 150 | 289 |
| V | 84 | 101 | 185 |
| VI | 16 | 15 | 31 |
| N/A | 3446 | 3093 | 6539 |
| Unknown | 307 | 490 | 797 |
| Modality |  |  |  |
| Contrast-enhanced X-Ray | 158 | 189 | 347 |
| CT | 742 | 667 | 1409 |
| Dermoscopy | 8 | 16 | 24 |
| Endoscopy | 220 | 244 | 464 |
| Fundoscopy | 55 | 42 | 97 |
| Microscopy | 665 | 706 | 1371 |
| MRI | 277 | 265 | 542 |
| Nuclear | 34 | 41 | 75 |
| OCT | 18 | 13 | 31 |
| Other | 6 | 7 | 13 |
| Photo | 1578 | 1413 | 2991 |
| Ultrasound | 226 | 213 | 439 |
| X-Ray | 481 | 442 | 923 |
| Unknown | 2 | 10 | 12 |
| Body region |  |  |  |
| Abdomen | 950 | 584 | 1534 |
| Head and neck | 1118 | 705 | 1823 |
| Lower extremity | 348 | 257 | 605 |
| Multiple | 234 | 103 | 337 |
| Thorax | 660 | 375 | 1035 |
| Upper extremity | 271 | 168 | 439 |
| N/A | 868 | 2027 | 2895 |
| Unknown | 21 | 49 | 70 |
| Speciality |  |  |  |
| Allergology | 0 | 1 | 1 |
| Anatomy | 3 | 2 | 5 |
| Anesthesiology | 27 | 13 | 40 |
| Cardiac surgery | 0 | 32 | 32 |
| Cardiology | 207 | 178 | 385 |
| Critical care | 26 | 9 | 35 |
| Dentistry | 326 | 194 | 520 |
| Dermatology | 326 | 308 | 634 |
| Emergency medicine | 0 | 6 | 6 |
| Endocrinology | 87 | 28 | 115 |
| Forensic medicine | 3 | 5 | 8 |
| Gastroenterology | 347 | 230 | 577 |
| General surgery | 258 | 377 | 635 |
| Genetics | 10 | 13 | 23 |
| Hematology | 40 | 47 | 87 |
| Hepatology | 0 | 1 | 1 |
| Immunology | 0 | 2 | 2 |
| Infectious diseases | 120 | 195 | 315 |
| Internal medicine | 13 | 20 | 33 |
| Microbiology | 0 | 15 | 15 |
| Neonatology | 0 | 3 | 3 |
| Nephrology | 52 | 54 | 106 |
| Neurology | 159 | 116 | 275 |
| Neurosurgery | 70 | 133 | 203 |
| Obstetrics and gynecology | 115 | 137 | 252 |
| Oncology | 512 | 418 | 930 |
| Ophthalmology | 201 | 173 | 374 |
| Oral and maxillofacial surgery | 73 | 155 | 228 |
| Orthopedics | 508 | 428 | 936 |
| Otorhinolaryngology | 123 | 119 | 242 |
| Parasitology | 0 | 1 | 1 |
| Pathology | 26 | 41 | 67 |
| Pediatric cardiology | 0 | 6 | 6 |
| Pediatrics | 107 | 113 | 220 |
| Physiology | 0 | 1 | 1 |
| Plastic surgery | 97 | 109 | 206 |
| Podiatry | 0 | 4 | 4 |
| Proctology | 6 | 3 | 9 |
| Psychiatry | 3 | 0 | 3 |
| Pulmonology | 187 | 149 | 336 |
| Radiology | 80 | 64 | 144 |
| Radiotherapy | 0 | 2 | 2 |
| Rehabilitation medicine | 15 | 6 | 21 |
| Rheumatology | 34 | 19 | 53 |
| Sports medicine | 6 | 7 | 13 |
| Thoracic surgery | 55 | 49 | 104 |
| Toxicology | 0 | 5 | 5 |
| Traditional medicine | 0 | 1 | 1 |
| Urology | 160 | 139 | 299 |
| Vascular surgery | 80 | 105 | 185 |
| N/A | 0 | 31 | 31 |
| Unknown | 8 | 1 | 9 |

## SUPPLEMENTARY

**Supplementary Fig 1:**
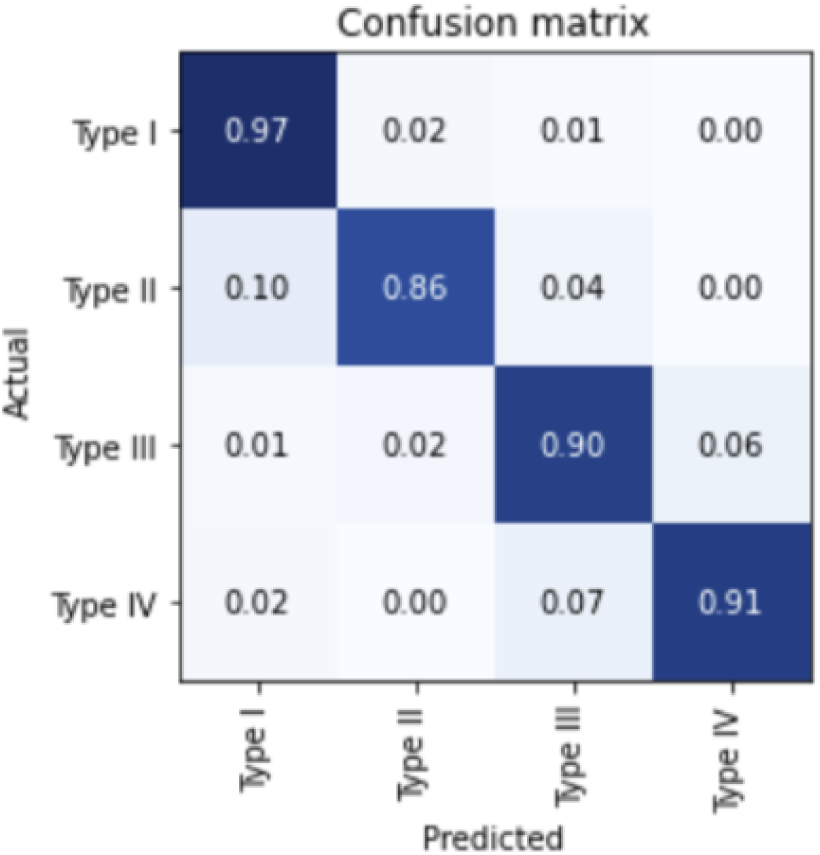
Normalized confusion matrix in the validation cohort (n=1000) for the Image Type CNN for classifying four different image types. Type I (single medical image), Type II (multiple medical images), Type III (one or multiple medical images combined with abstract images), Type IV (one or multiple abstract images). We manually labeled images (n=10000) from the PMCMID according to their image type. We split this dataset 90%/10% into a training and validation cohort and trained a ResNet34 Model on the training cohort. We verified the performance in the validation cohort. A high classification performance could be observed across all image types.

**Supplementary Fig 2:**
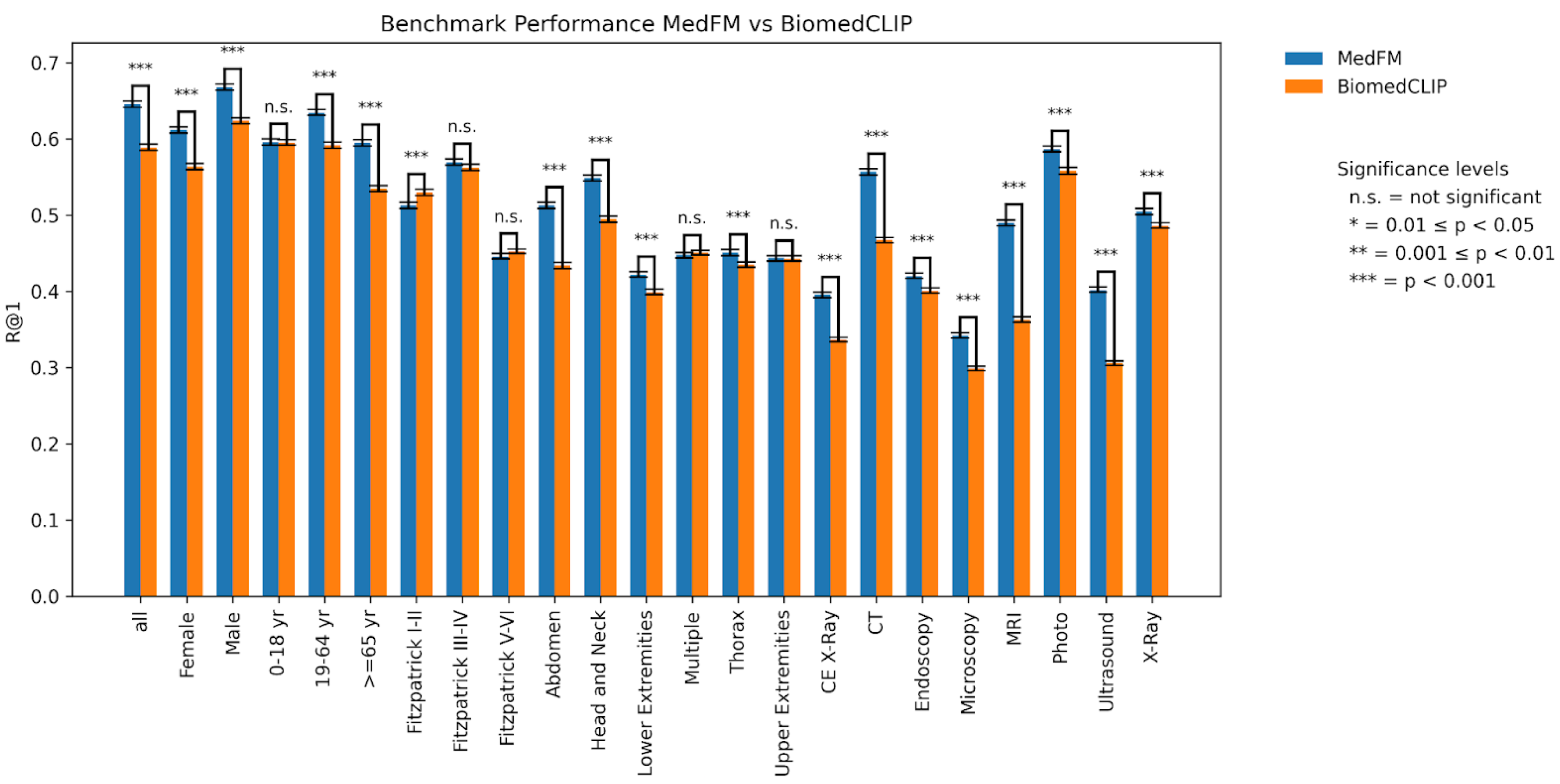
Comparison of MedFM and BiomedCLIP performance. MedFM outperformed BiomedCLIP, achieving an R@1 of 0.65 across all images compared to 0.59 for BiomedCLIP (p < 0.001). Significant improvements were observed in 16 of 22 sub-cohorts, with the highest improvement of 0.127 in the MRI sub-cohort and an average improvement of 0.053 (range: 0.016 to 0.127; p < 0.001).

**Supplementary Fig 3:**
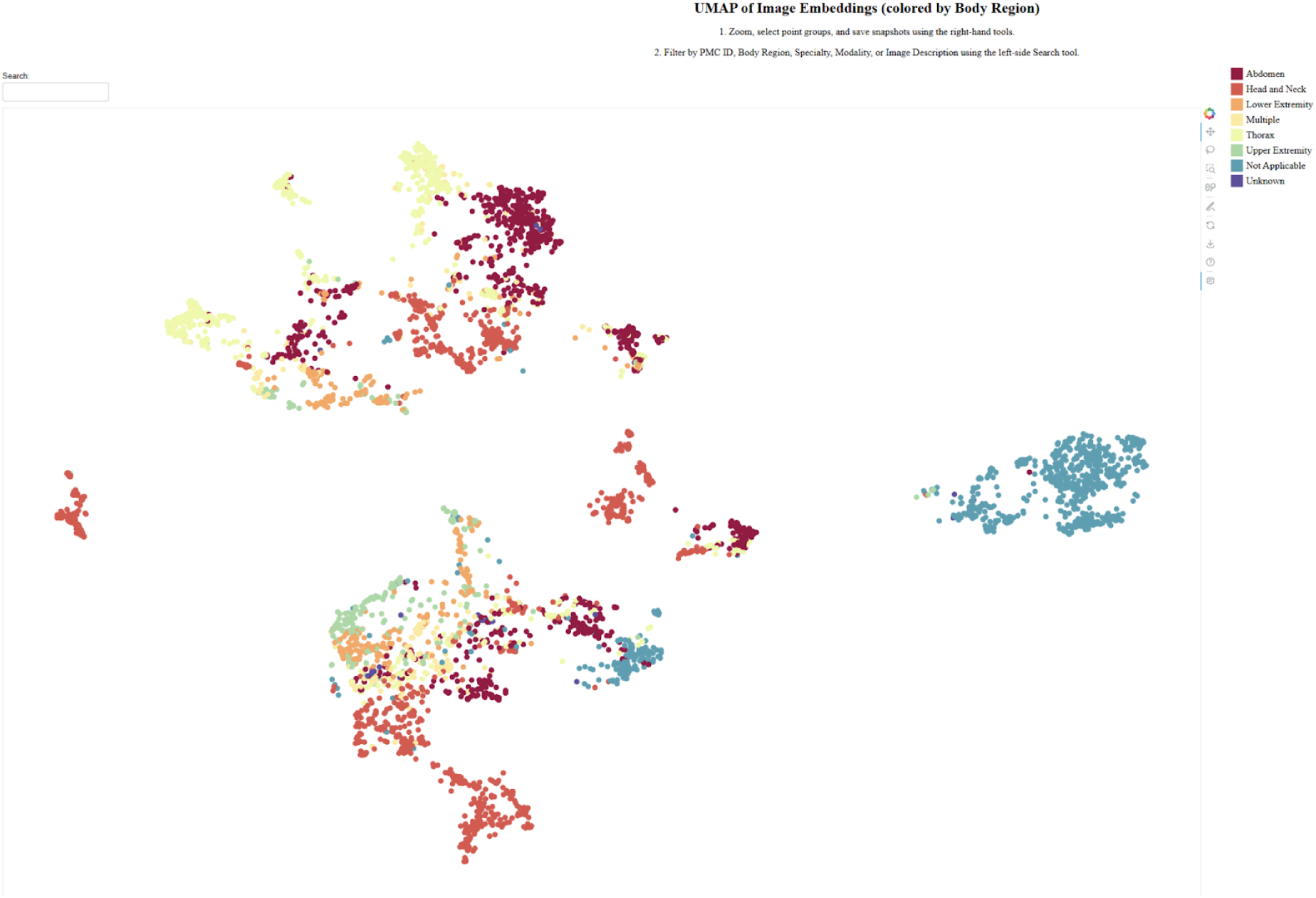
UMAP of Image Embeddings colored by Body Region: The UMAP visualization revealed distinct macro-clusters for certain body parts, such as Head and Neck (center-left), Thorax (top-left), Abdomen (top-right), and Lower Extremity (bottom). Additionally, it identifies several micro-clusters within these macro-clusters, including Lower Extremity splitting into three distant micro-clusters, Head and Neck into four, Abdomen into six, and Thorax into four micro-clusters. These patterns suggest the presence of shared visual features among different modalities that MedFM had identified and encoded into closer embeddings, reflecting potential modality-invariant characteristics.

**Supplementary Fig 4:**
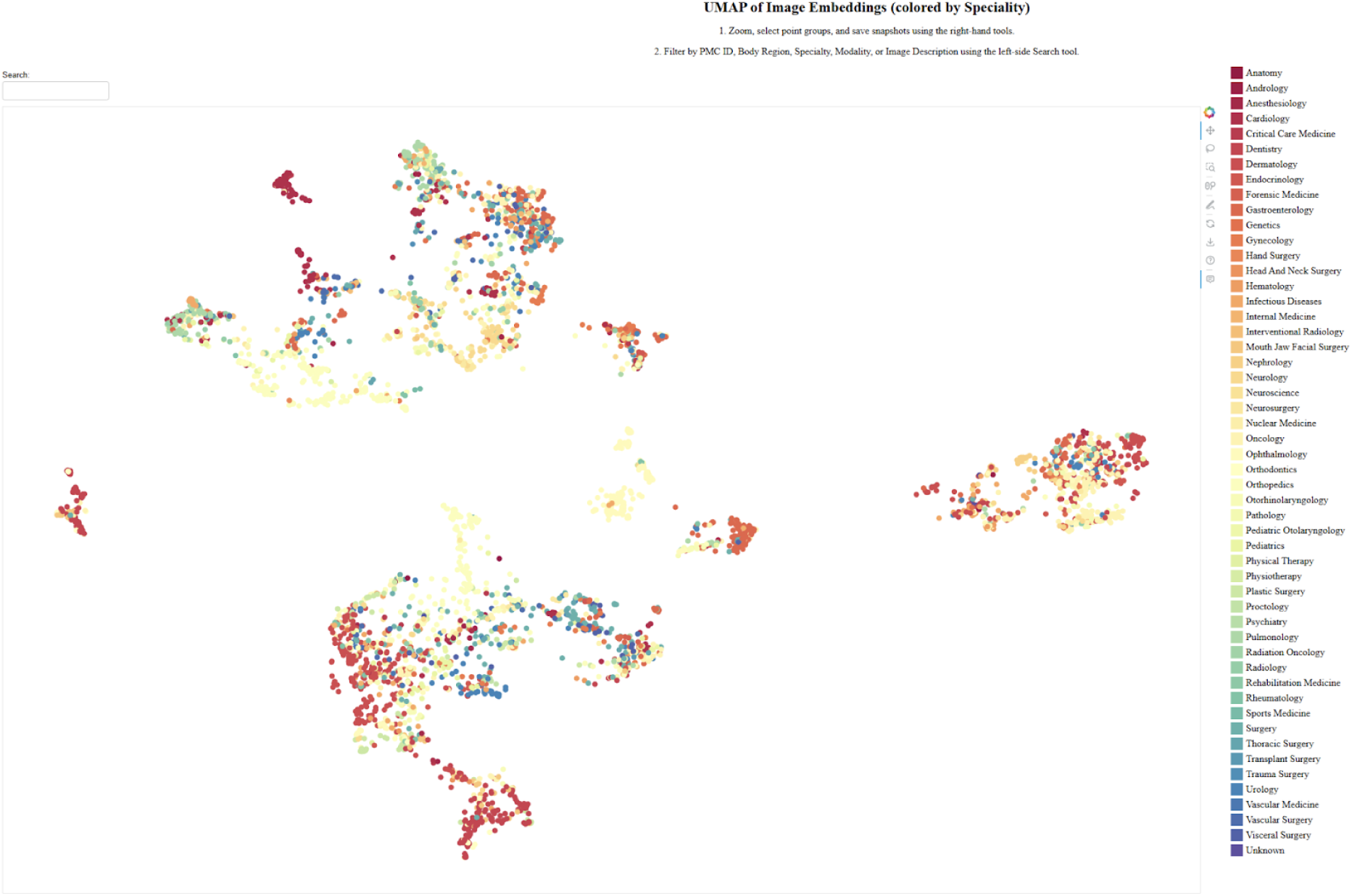
UMAP of Image Embeddings colored by Speciality: Despite the large number of specialties, a few distinct clusters with clear topological patterns are still observable. This is likely due to the significant similarities between images belonging to the same specialty and, conversely, the reuse of a single image across multiple specialties.

**Supplementary Fig 5:**
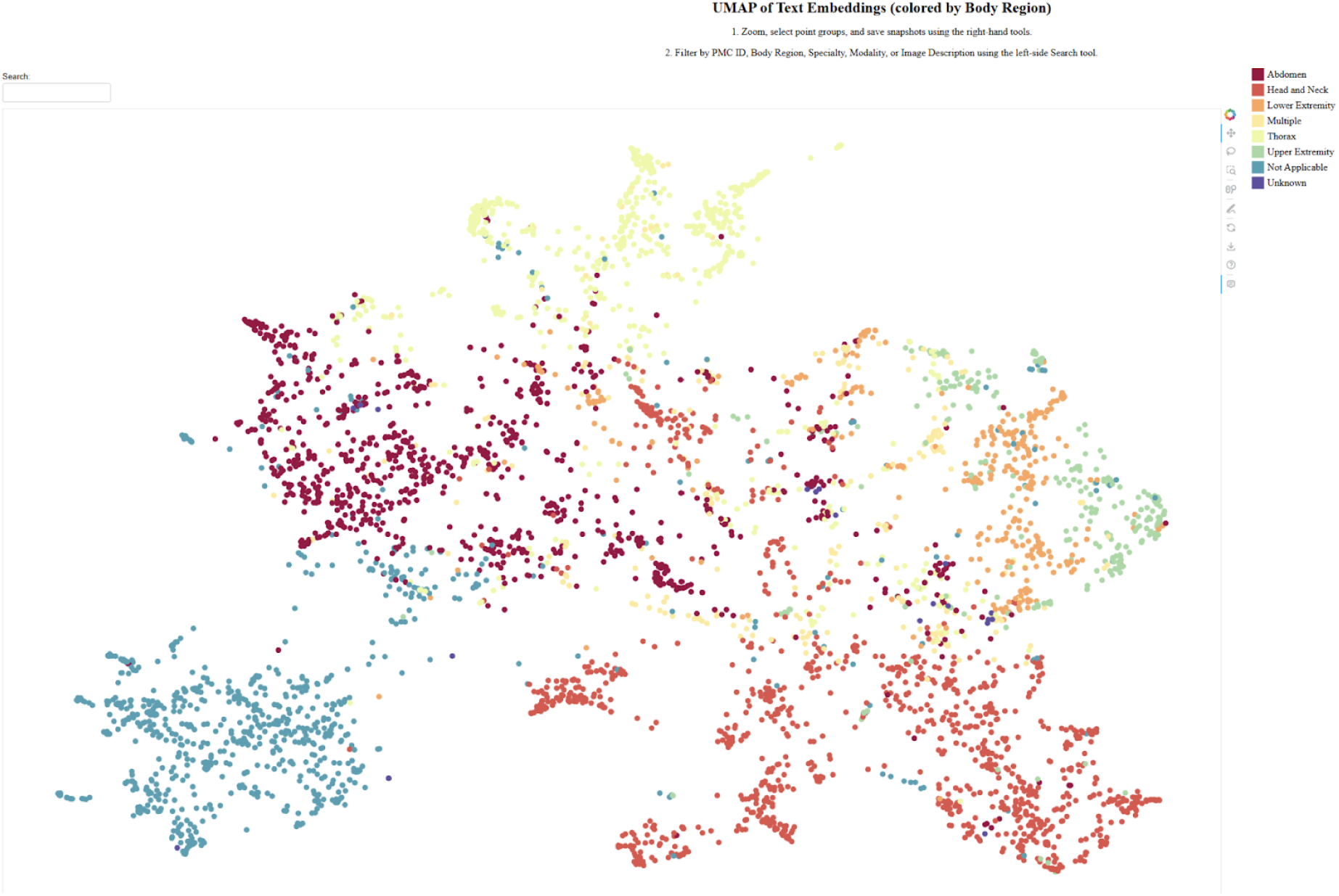
UMAP of Text Embeddings colored by Body Region: In the case of text embeddings, it is more challenging to achieve clear separation between clusters compared to image embeddings. This can likely be attributed to the richness of text descriptions, which often include numerous medical terms that are difficult to associate with specific body regions. Nevertheless, distinct clusters are still observable for Head and Neck (bottom-right), Abdomen (top-left), Thorax (top), and Upper Extremity (right). Additionally, multiple micro-clusters of Thorax appear interspersed among other clusters.

**Supplementary Fig 6:**
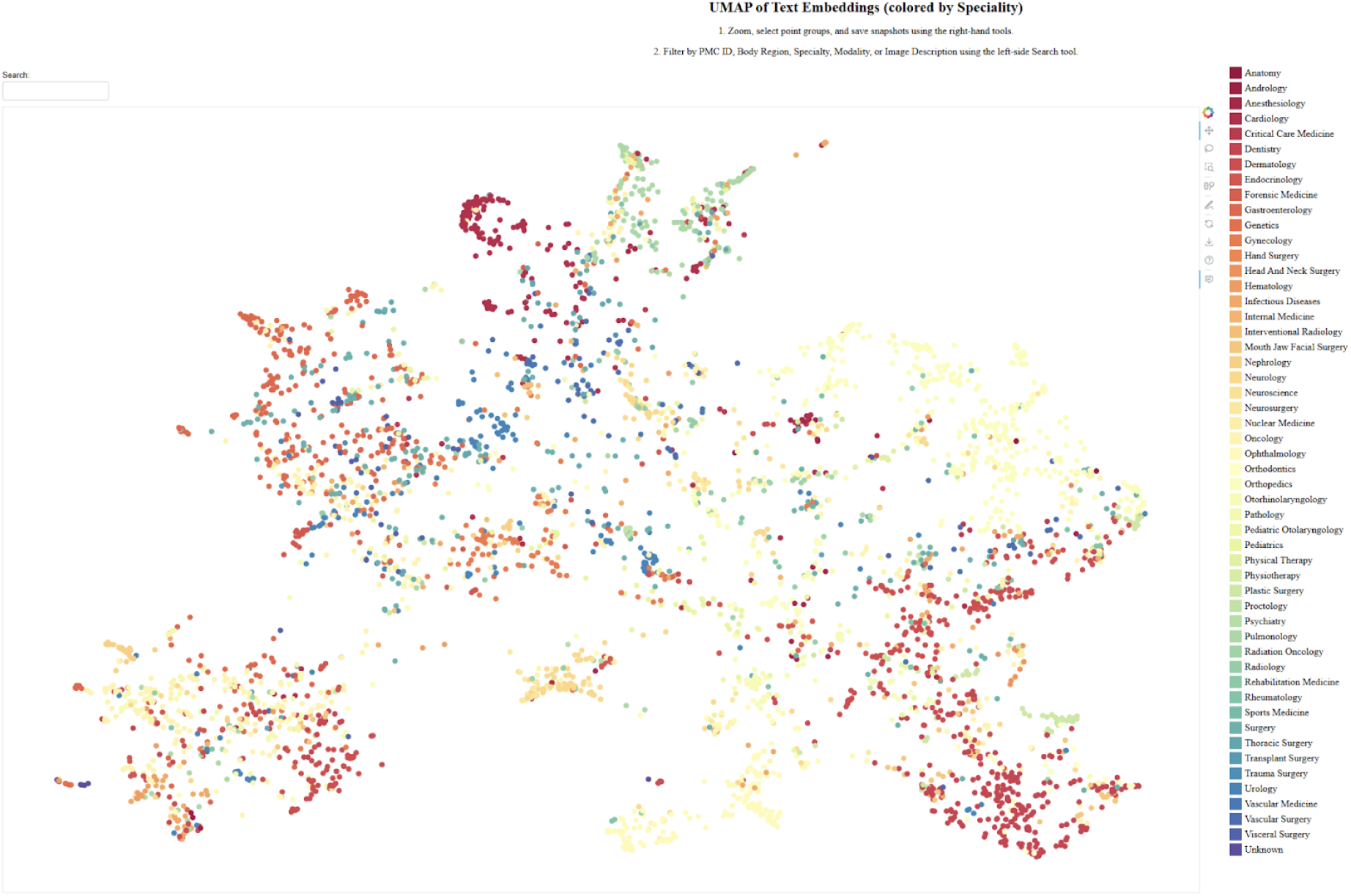
UMAP of Text Embeddings colored by Speciality. Similar to the case of image embeddings, the presence of numerous specialties makes it challenging to identify distinct macro-clusters.

**Supplementary Fig 7:**
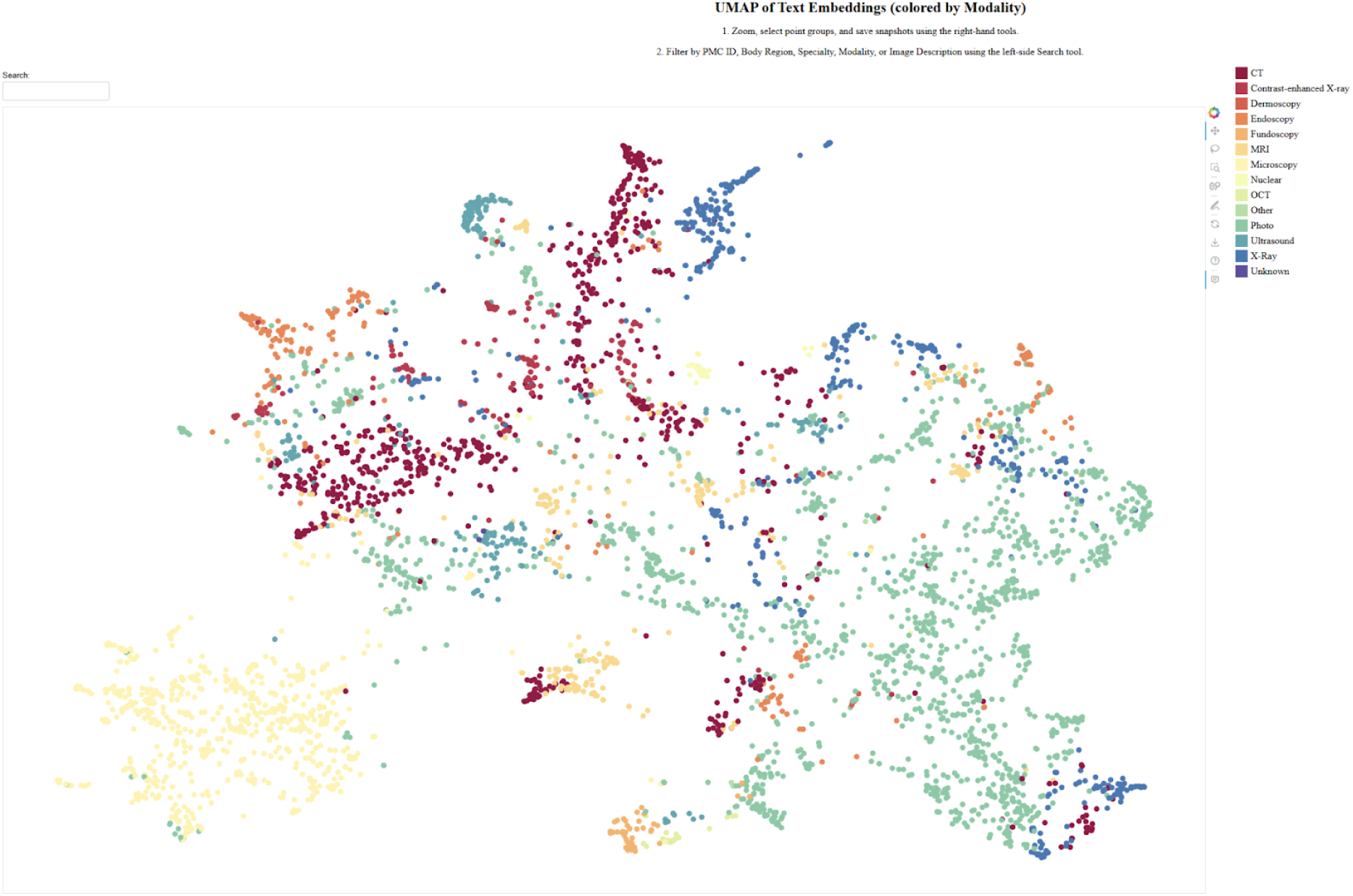
UMAP of Text Embeddings colored by Image Modality. The UMAP visualization of text embeddings, colored by modality, reveals distinct macro and micro clusters, along with their respective positions and topologies. Ultrasound and Dermoscopy clusters are distinct and well-separated, indicating unique textual patterns, while CT, MRI, and X-ray clusters partially overlap, reflecting shared features due to clinical similarities. MRI shows the largest variability, capturing diverse descriptions, while sparse peripheral clusters (e.g., Contrast-enhanced X-ray) and scattered groups (e.g., Fundoscopy, Endoscopy) indicate less common or ambiguous embeddings. The visualization highlights modality-specific behaviors and opportunities for improving separability through further analysis.

**Supplementary Fig 8:**
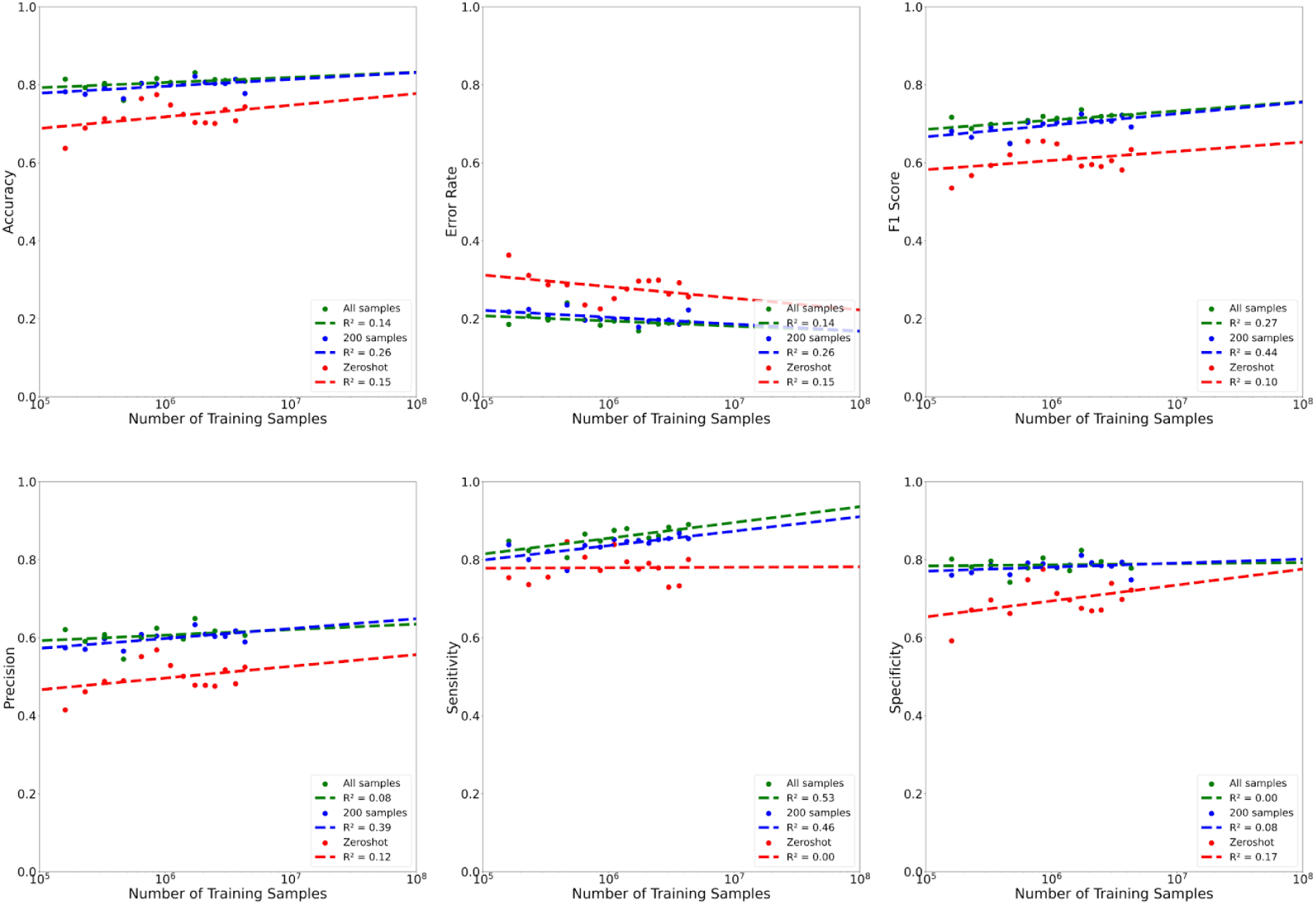
Scaling plots of various performance metrics versus the number of PMCMID training images in the pneumonia classification use case.

**Supplementary Fig 9:**
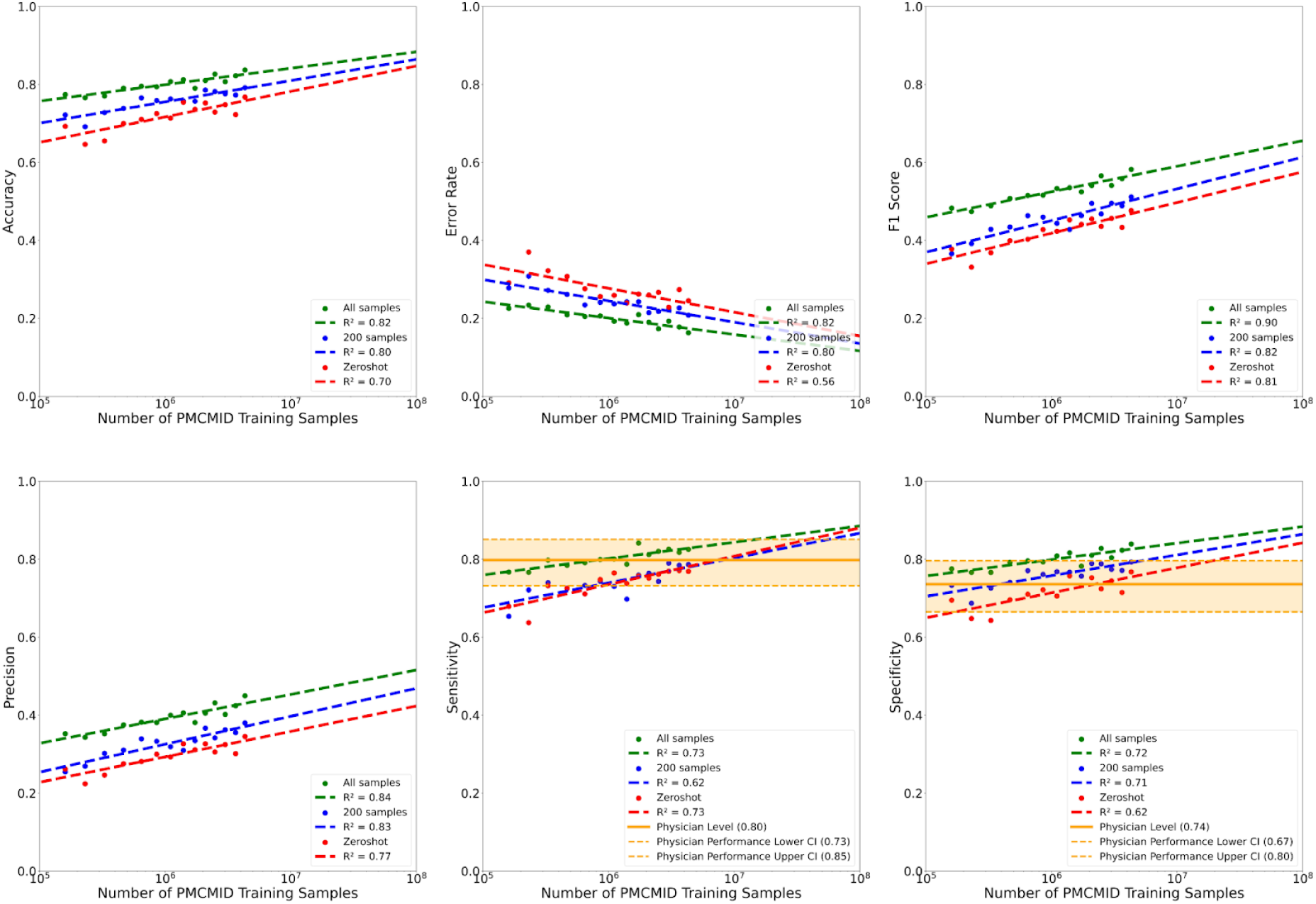
Scaling plots of Accuracy, Error Rate, F1 Score, Precision, Sensitivity, and Specificity versus the number of PMCMID training images for skin cancer classification.

**Supplementary Fig 10:**
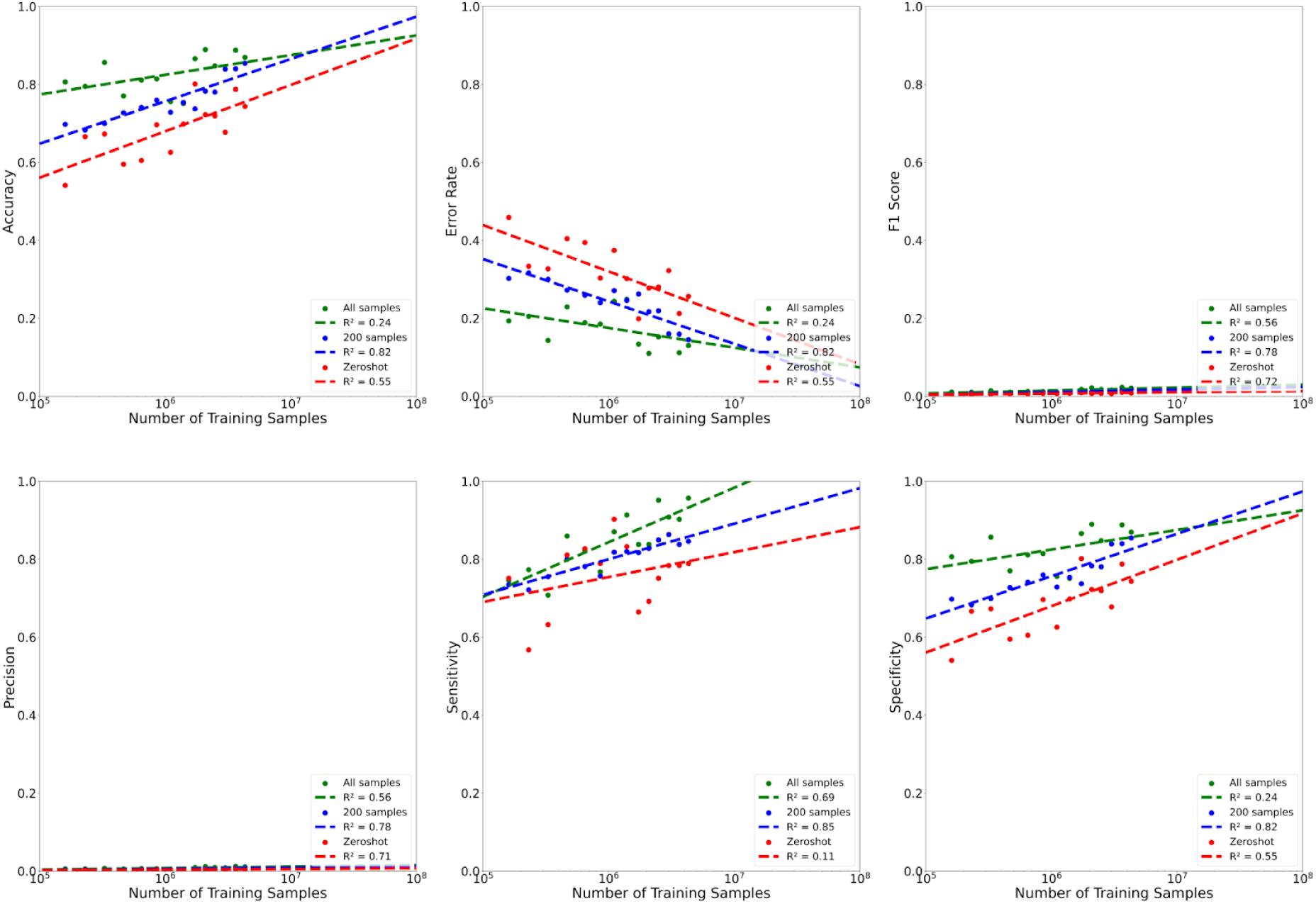
Scaling plots of Accuracy, Error Rate, F1 Score, Precision, Sensitivity, and Specifity versus the number of PMCMID training images for mpox classification.

**Supplementary Fig 11:**
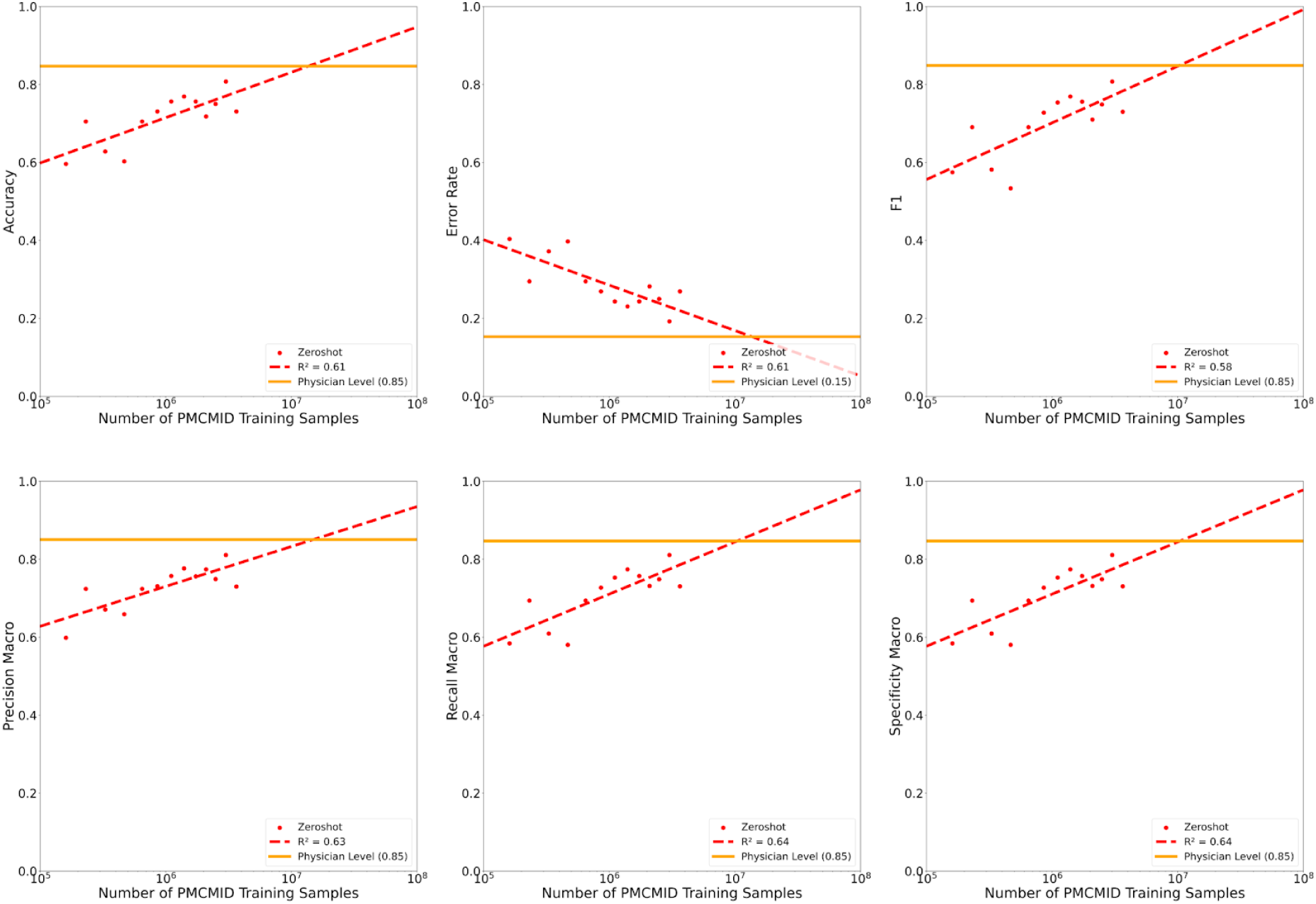
Scaling plots of Accuracy, Error Rate, F1 Score, Precision, Sensitivity, and Specifity versus the number of PMCMID training images for lung cancer subtype classification.

## Supplementary Tables

**Supplementary Table 1:** Performance of the MedFM measured as the average R@1 and 95% confidence intervals for ranking diagnoses in 50 random medical images sampled from all images or subcohorts of the PMCMID test set.

|  |  | Mean | 95%CI<br>(Lower) | 94%C<br>(Upper) |
| --- | --- | --- | --- | --- |
| All images |  | 0.65 | 0.64 | 0.67 |
|  | Subcohort |  |  |  |
| Sex | Female | 0.62 | 0.61 | 0.63 |
|  | Male | 0.67 | 0.66 | 0.68 |
| Age | 0-18 yr | 0.60 | 0.59 | 0.61 |
|  | 19-64 yr | 0.63 | 0.61 | 0.64 |
|  | >=65 yr | 0.61 | 0.60 | 0.62 |
| Skin Tone | Fitzpatrick I-II | 0.51 | 0.50 | 0.52 |
|  | Fitzpatrick III-IV | 0.58 | 0.56 | 0.59 |
|  | Fitzpatrick V-VI | 0.44 | 0.43 | 0.45 |
| Body Region | Upper Extremities | 0.44 | 0.42 | 0.45 |
|  | Lower Extremities | 0.42 | 0.40 | 0.43 |
|  | Head and Neck | 0.55 | 0.54 | 0.57 |
|  | Multiple | 0.44 | 0.43 | 0.45 |
|  | Abdomen | 0.51 | 0.50 | 0.53 |
|  | Thorax | 0.46 | 0.45 | 0.47 |
| Image Modality | MRI | 0.48 | 0.47 | 0.50 |
|  | CT | 0.54 | 0.53 | 0.56 |
|  | CE X-Ray | 0.40 | 0.39 | 0.41 |
|  | X-Ray | 0.50 | 0.49 | 0.52 |
|  | Microscopy | 0.35 | 0.34 | 0.36 |
|  | Ultrasound | 0.41 | 0.40 | 0.42 |
|  | Photo | 0.60 | 0.59 | 0.61 |
|  | Endoscopy | 0.42 | 0.41 | 0.43 |

**Supplementary Table 2:** Datasets and data sizes used for training and testing in the Pneumonia/Radiology evaluation use case.

|  |  | CheXpert |  |  | PadChest |  | MIMIC |  | RSNA |  | Total |
| --- | --- | --- | --- | --- | --- | --- | --- | --- | --- | --- | --- |
|  |  | Train | Test | Excluded | Test | Excluded | Test | Excluded | Test | Excluded |  |
| age | 0-18 y.o. | 34 | 2 | 737 | 1059 | 6585 | 0 | 0 | 0 | 0 | 8417 |
|  | 19-64 y.o. | 3031 | 459 | 121521 | 13335 | 66735 | 0 | 0 | 0 | 0 | 205081 |
|  | >=65 y.o | 2820 | 277 | 95435 | 12038 | 61070 | 0 | 0 | 0 | 0 | 171640 |
|  | Unknown | 0 | 0 | 0 | 0 | 6 | 2004 | 375106 | 30227 | 0 | 407343 |
| sex | Female | 2396 | 332 | 88449 | 13773 | 66128 | 0 | 0 | 0 | 0 | 171078 |
|  | Male | 3489 | 406 | 129243 | 12656 | 68249 | 0 | 0 | 0 | 0 | 214043 |
|  | Unknown | 0 | 0 | 1 | 3 | 19 | 2004 | 375106 | 30227 | 0 | 407360 |
| projection | AP | 3845 | 566 | 157745 | 2471 | 16430 | 1074 | 146099 | 30227 | 0 | 358457 |
|  | PA | 2040 | 172 | 27380 | 23961 | 67752 | 930 | 95231 | 0 | 0 | 217466 |
|  | L | 0 | 0 | 32419 | 0 | 49573 | 0 | 82853 | 0 | 0 | 164845 |
|  | Lateral Left | 0 | 0 | 16 | 0 | 0 | 0 | 35133 | 0 | 0 | 35149 |
|  | Other | 0 | 0 | 133 | 0 | 641 | 0 | 15790 | 0 | 0 | 16564 |
| pneumonia | no | 2148 | 718 | 197186 | 25367 | 131987 | 1539 | 320199 | 20672 | 0 | 699816 |
|  | yes | 3737 | 20 | 1121 | 1065 | 2409 | 465 | 25757 | 9555 | 0 | 44129 |
|  | uncertain | 0 | 0 | 19386 | 0 | 0 | 0 | 29150 | 0 | 0 | 48536 |
| physician annotated | yes | 5885 | 738 | 0 | 26432 | 0 | 2004 | 0 | 30227 | 0 | 65286 |
|  | no | 0 | 0 | 217693 | 0 | 134396 | 0 | 375106 | 0 | 0 | 727195 |

**Supplementary Table 3:** Medical terms for pneumonia classification in CRX.

| # | Pneumonia Classification Terms |
| --- | --- |
| 1 | pneumonia |
| 2 | pneumonia with airspace opacification |
| 3 | pneumonia with air bronchograms |
| 4 | pneumonia with cavitation |
| 5 | pneumonia with pleural collection |
| 6 | aspiration pneumonia |
| 7 | atypical pneumonia |
| 8 | bilateral pneumonia |
| 9 | bronchopneumonia |
| 10 | cavitating pneumonia |
| 11 | interstitial pneumonia |
| 12 | lobar pneumonia |
| 13 | viral pneumonia |
| 14 | air bronchogram |
| 15 | consolidation |
| 16 | ground-glass opacity |
| 17 | infiltrate |
| 18 | interstitial pattern |
| 19 | opacity |
| 20 | pleural effusions |
| 21 | lung opacity |
| 22 | healthy |
| 23 | healthy lung |
| 24 | no finding |
| 25 | normal lung |
| 26 | sharp costophrenic angles |

**Supplementary Table 4:** Pneumonia - Zero-shot.

| Model<br>(MedFM<br>_YYYY) | Training<br>Images<br>(n) | AUC | Accuracy | Balanced<br>Accuracy | Error<br>Rate | Sensitivity | Specificity | Precision | F1 Score | R@1 |
| --- | --- | --- | --- | --- | --- | --- | --- | --- | --- | --- |
| 2010 | 160766 | 0.725 | 0.637 | 0.673 | 0.363 | 0.754 | 0.592 | 0.415 | 0.535 | 0.224 |
| 2011 | 231351 | 0.762 | 0.689 | 0.703 | 0.311 | 0.736 | 0.671 | 0.461 | 0.567 | 0.256 |
| 2012 | 330034 | 0.789 | 0.713 | 0.726 | 0.287 | 0.755 | 0.697 | 0.488 | 0.593 | 0.277 |
| 2013 | 468100 | 0.811 | 0.713 | 0.754 | 0.287 | 0.846 | 0.662 | 0.49 | 0.62 | 0.324 |
| 2014 | 648173 | 0.841 | 0.765 | 0.778 | 0.235 | 0.806 | 0.749 | 0.551 | 0.655 | 0.347 |
| 2015 | 858263 | 0.843 | 0.775 | 0.774 | 0.225 | 0.773 | 0.776 | 0.569 | 0.655 | 0.375 |
| 2016 | 1106765 | 0.843 | 0.748 | 0.776 | 0.252 | 0.839 | 0.713 | 0.529 | 0.649 | 0.402 |
| 2017 | 1397964 | 0.809 | 0.724 | 0.746 | 0.276 | 0.795 | 0.697 | 0.501 | 0.615 | 0.401 |
| 2018 | 1734575 | 0.788 | 0.703 | 0.726 | 0.297 | 0.776 | 0.675 | 0.478 | 0.592 | 0.43 |
| 2019 | 2095551 | 0.787 | 0.702 | 0.73 | 0.298 | 0.791 | 0.669 | 0.478 | 0.596 | 0.426 |
| 2020 | 2493887 | 0.78 | 0.701 | 0.725 | 0.299 | 0.779 | 0.671 | 0.476 | 0.59 | 0.473 |
| 2021 | 3014957 | 0.788 | 0.737 | 0.734 | 0.263 | 0.729 | 0.739 | 0.517 | 0.605 | 0.487 |
| 2022 | 3654626 | 0.771 | 0.708 | 0.716 | 0.292 | 0.733 | 0.698 | 0.482 | 0.582 | 0.492 |
| 2023 | 4326353 | 0.822 | 0.744 | 0.761 | 0.256 | 0.801 | 0.722 | 0.525 | 0.634 | 0.507 |

**Supplementary Table 5:**
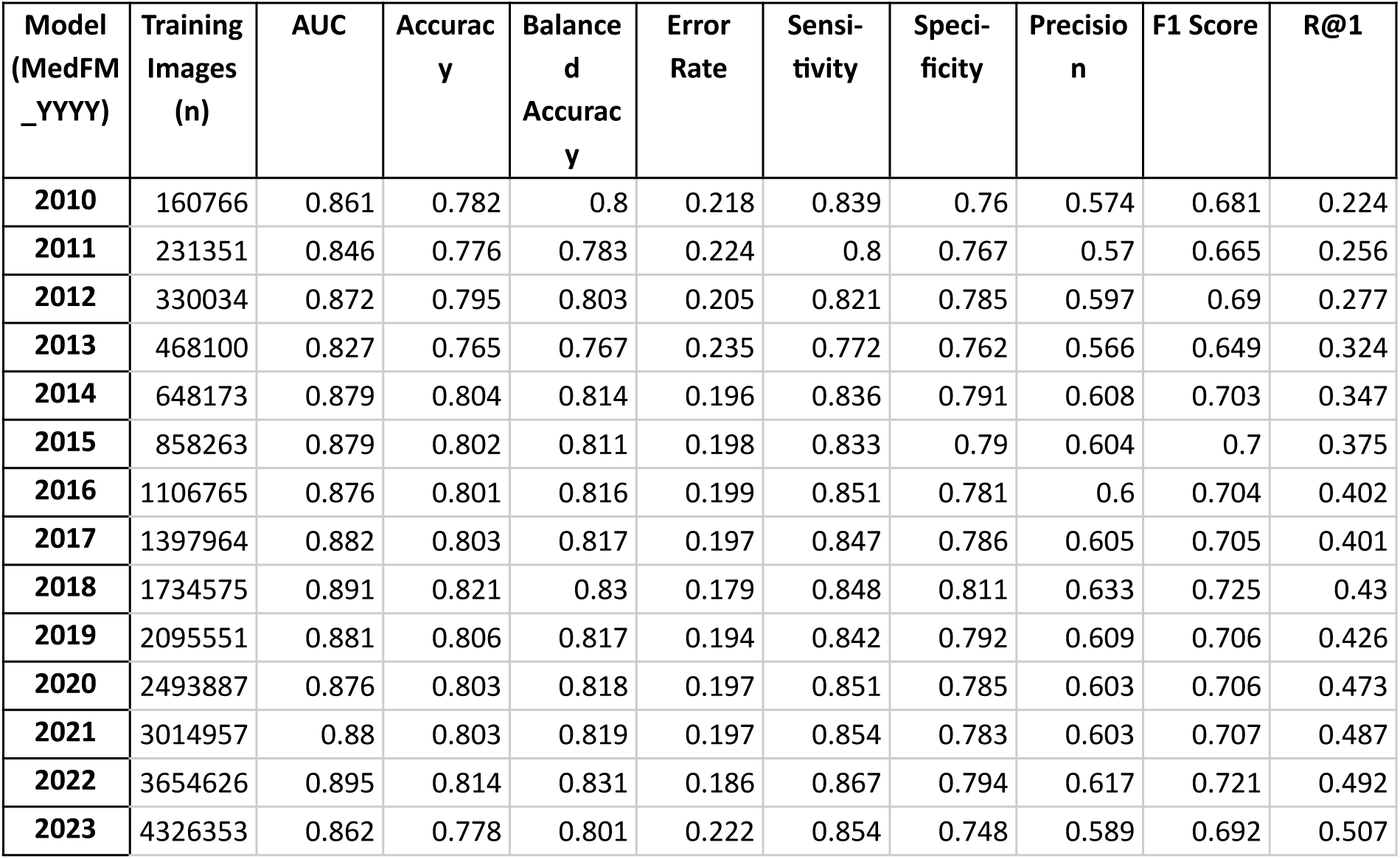
Pneumonia- 200 Samples.

**Supplementary Table 6:**
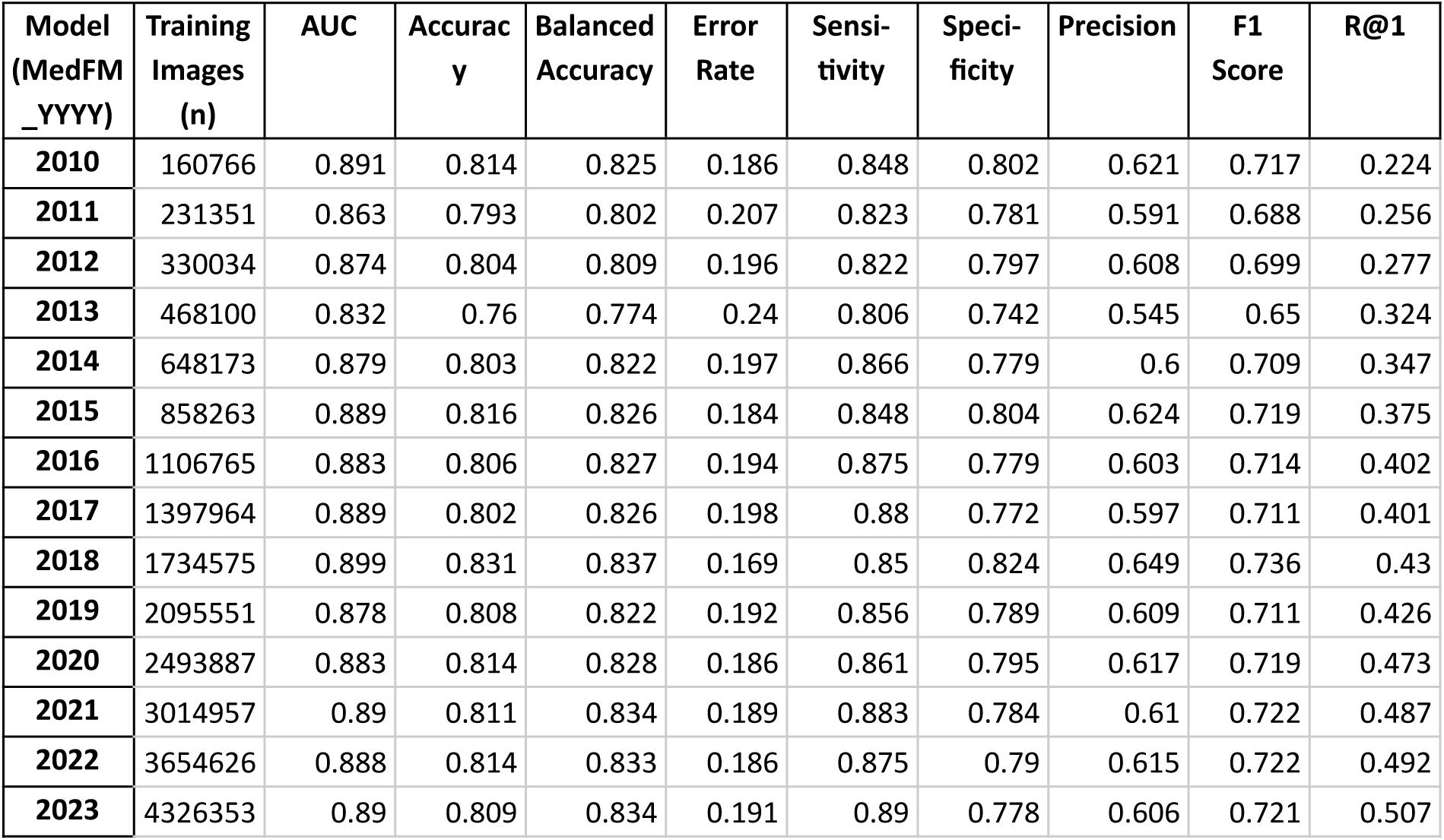
Pneumonia- All Samples.

| Model<br>(MedFM<br>_YYYY) | Training<br>Images<br>(n) | AUC | Accuracy | Balanced<br>Accuracy | Error<br>Rate | Sensitivity | Specificity | Precision | F1<br>Score | R@1 |
| --- | --- | --- | --- | --- | --- | --- | --- | --- | --- | --- |
| 2010 | 160766 | 0.891 | 0.814 | 0.825 | 0.186 | 0.848 | 0.802 | 0.621 | 0.717 | 0.224 |
| 2011 | 231351 | 0.863 | 0.793 | 0.802 | 0.207 | 0.823 | 0.781 | 0.591 | 0.688 | 0.256 |
| 2012 | 330034 | 0.874 | 0.804 | 0.809 | 0.196 | 0.822 | 0.797 | 0.608 | 0.699 | 0.277 |
| 2013 | 468100 | 0.832 | 0.76 | 0.774 | 0.24 | 0.806 | 0.742 | 0.545 | 0.65 | 0.324 |
| 2014 | 648173 | 0.879 | 0.803 | 0.822 | 0.197 | 0.866 | 0.779 | 0.6 | 0.709 | 0.347 |
| 2015 | 858263 | 0.889 | 0.816 | 0.826 | 0.184 | 0.848 | 0.804 | 0.624 | 0.719 | 0.375 |
| 2016 | 1106765 | 0.883 | 0.806 | 0.827 | 0.194 | 0.875 | 0.779 | 0.603 | 0.714 | 0.402 |
| 2017 | 1397964 | 0.889 | 0.802 | 0.826 | 0.198 | 0.88 | 0.772 | 0.597 | 0.711 | 0.401 |
| 2018 | 1734575 | 0.899 | 0.831 | 0.837 | 0.169 | 0.85 | 0.824 | 0.649 | 0.736 | 0.43 |
| 2019 | 2095551 | 0.878 | 0.808 | 0.822 | 0.192 | 0.856 | 0.789 | 0.609 | 0.711 | 0.426 |
| 2020 | 2493887 | 0.883 | 0.814 | 0.828 | 0.186 | 0.861 | 0.795 | 0.617 | 0.719 | 0.473 |
| 2021 | 3014957 | 0.89 | 0.811 | 0.834 | 0.189 | 0.883 | 0.784 | 0.61 | 0.722 | 0.487 |
| 2022 | 3654626 | 0.888 | 0.814 | 0.833 | 0.186 | 0.875 | 0.79 | 0.615 | 0.722 | 0.492 |
| 2023 | 4326353 | 0.89 | 0.809 | 0.834 | 0.191 | 0.89 | 0.778 | 0.606 | 0.721 | 0.507 |

**Supplementary Table 7:** Datasets and data sizes used for training and testing in the Skin cancer evaluation use case.

| Category | Subcategory | DermNetNZ | Danderm | Dermis | DermNet | Esteva | Fitzpatrick 17k | Hellenic | PAD UFES | Total |
| --- | --- | --- | --- | --- | --- | --- | --- | --- | --- | --- |
| Excluded |  | 1,973 | 5 | 0 | 0 | 45440 | 52 | 48 | 0 | 47,518 |
| Included |  | 12045 | 3432 | 6,589 | 19,289 | 75730 | 16525 | 2614 | 2,298 | 138522 |
| Training |  | 0 | 0 | 0 | 0 | 75730 | 0 | 0 | 0 | 75730 |
| Testing |  | 12045 | 3432 | 6589 | 19289 | 0 | 16525 | 2614 | 2298 | 62792 |
| age | 0-18 y.o. | 0 | 0 | 509 | 0 | 0 | 0 | 0 | 40 | 549 |
|  | 19-64 y.o. | 0 | 0 | 1932 | 0 | 0 | 0 | 0 | 1251 | 3183 |
|  | >=65 y.o | 0 | 0 | 572 | 0 | 0 | 0 | 0 | 1007 | 1579 |
|  | Unknown | 12045 | 3432 | 3576 | 19289 | 75730 | 16525 | 2614 | 0 | 133211 |
| sex | Female | 0 | 0 | 2520 | 0 | 0 | 0 | 0 | 753 | 3273 |
|  | Male | 0 | 0 | 2593 | 0 | 0 | 0 | 0 | 741 | 3334 |
|  | Unknown | 12045 | 3432 | 1476 | 19289 | 75730 | 16525 | 2614 | 804 | 131915 |
| malignant | yes | 976 | 187 | 771 | 1583 | 6953 | 2036 | 223 | 1819 | 14548 |
|  | no | 5754 | 2363 | 5058 | 17256 | 68174 | 13857 | 2181 | 479 | 115122 |
|  | Unknown | 5315 | 882 | 760 | 450 | 603 | 632 | 210 | 0 | 8852 |
| skin tone (Fitzpatrick) | Fitzpatrick I | 0 | 0 | 0 | 0 | 0 | 2941 | 0 | 153 | 3094 |
|  | Fitzpatrick II | 0 | 0 | 0 | 0 | 0 | 4796 | 0 | 876 | 5672 |
|  | Fitzpatrick III | 0 | 0 | 0 | 0 | 0 | 3296 | 0 | 392 | 3688 |
|  | Fitzpatrick IV | 0 | 0 | 0 | 0 | 0 | 2775 | 0 | 62 | 2837 |
|  | Fitzpatrick V | 0 | 0 | 0 | 0 | 0 | 1527 | 0 | 10 | 1537 |
|  | Fitzpatrick VI | 0 | 0 | 0 | 0 | 0 | 628 | 0 | 1 | 629 |
|  | Unknown | 12045 | 3432 | 6589 | 19289 | 75730 | 562 | 2614 | 804 | 121065 |
| body region | Head | 0 | 0 | 1443 | 0 | 0 | 0 | 0 | 842 | 2285 |
|  | Neck | 0 | 0 | 96 | 0 | 0 | 0 | 0 | 93 | 189 |
|  | Torso | 0 | 0 | 705 | 0 | 0 | 0 | 0 | 564 | 1269 |
|  | Upper extremity | 0 | 0 | 916 | 0 | 0 | 0 | 0 | 710 | 1626 |
|  | Lower extremity | 0 | 0 | 813 | 0 | 0 | 0 | 0 | 89 | 902 |
|  | Anogenital | 0 | 0 | 223 | 0 | 0 | 0 | 0 | 0 | 223 |
|  | Multiple Body Regions | 0 | 0 | 110 | 0 | 0 | 0 | 0 | 0 | 110 |
|  | Unknown | 12045 | 3432 | 2283 | 19289 | 75730 | 16525 | 2614 | 0 | 131918 |

**Supplementary Table 8:** Medical terms for skin cancer classification in clinical photos.

| # | Medical terms for skin cancer |
| --- | --- |
| 1 | skin lesion with border irregularity |
| 2 | skin lesion with uniform coloration |
| 3 | skin lesion with color variation |
| 4 | skin lesion with smooth texture |
| 5 | skin lesion with ulceration |
| 6 | asymmetric skin lesion |
| 7 | bleeding skin lesion |
| 8 | skin cancer |
| 9 | cancer |
| 10 | malignant |
| 11 | malignant skin lesion |
| 12 | benign skin lesion |
| 13 | normal |
| 14 | hair in skin lesion |
| 15 | atypical skin lesion |
| 16 | pigmented skin lesion |
| 17 | normal skin |
| 18 | keratoacanthoma |
| 19 | erythema multiforme and fixed drug eruptions |
| 20 | seborrheic keratosis |
| 21 | rosacea |
| 22 | intermediate or malignant vascular tumors |
| 23 | scabies |
| 24 | bowens disease |
| 25 | eczema |
| 26 | melanocytic nevi |
| 27 | malignant melanoma |
| 28 | solar damage actinic keratosis |
| 29 | symmetric skin lesion |
| 30 | systemic carcinomas |
| 31 | psoriasis |
| 32 | actinic keratosis |
| 33 | herpes zoster |
| 34 | squamous cell carcinoma |
| 35 | cutaneous t cell lymphoma |
| 36 | acne vulgaris |
| 37 | lichen planus |
| 38 | basal cell carcinoma |

**Supplementary Table 9:** Skin Cancer - Zero-shot.

| Model<br>(MedFM<br>_YYYY) | Training<br>Images<br>(n) | AUC | Accuracy | Balanced<br>Accuracy | Error<br>Rate | Sensitivity | Specificity | Precision | F1<br>Score | R@1 |
| --- | --- | --- | --- | --- | --- | --- | --- | --- | --- | --- |
| 2010 | 160766 | 0.785 | 0.709 | 0.715 | 0.291 | 0.723 | 0.707 | 0.282 | 0.406 | 0.279 |
| 2011 | 231351 | 0.728 | 0.63 | 0.668 | 0.37 | 0.721 | 0.615 | 0.23 | 0.349 | 0.319 |
| 2012 | 330034 | 0.776 | 0.678 | 0.708 | 0.322 | 0.751 | 0.666 | 0.264 | 0.39 | 0.361 |
| 2013 | 468100 | 0.79 | 0.693 | 0.719 | 0.307 | 0.757 | 0.682 | 0.275 | 0.404 | 0.402 |
| 2014 | 648173 | 0.79 | 0.724 | 0.722 | 0.276 | 0.72 | 0.725 | 0.294 | 0.418 | 0.414 |
| 2015 | 858263 | 0.814 | 0.744 | 0.746 | 0.256 | 0.748 | 0.744 | 0.317 | 0.445 | 0.449 |
| 2016 | 1106765 | 0.817 | 0.741 | 0.747 | 0.259 | 0.755 | 0.739 | 0.316 | 0.445 | 0.474 |
| 2017 | 1397964 | 0.82 | 0.761 | 0.747 | 0.239 | 0.729 | 0.766 | 0.332 | 0.456 | 0.483 |
| 2018 | 1734575 | 0.819 | 0.738 | 0.747 | 0.262 | 0.759 | 0.734 | 0.313 | 0.443 | 0.506 |
| 2019 | 2095551 | 0.829 | 0.741 | 0.757 | 0.259 | 0.781 | 0.734 | 0.319 | 0.453 | 0.527 |
| 2020 | 2493887 | 0.809 | 0.733 | 0.737 | 0.267 | 0.743 | 0.732 | 0.306 | 0.434 | 0.525 |
| 2021 | 3014957 | 0.835 | 0.771 | 0.76 | 0.229 | 0.745 | 0.775 | 0.346 | 0.472 | 0.552 |
| 2022 | 3654626 | 0.819 | 0.726 | 0.745 | 0.274 | 0.77 | 0.719 | 0.304 | 0.436 | 0.573 |
| 2023 | 4326353 | 0.848 | 0.755 | 0.77 | 0.245 | 0.79 | 0.749 | 0.334 | 0.47 | 0.587 |

**Supplementary Table 10:** Skin Cancer - 200 Samples.

| Model<br>(MedFM<br>_YYYY) | Training<br>Images<br>(n) | AUC | Accuracy | Balanced<br>Accuracy | Error<br>Rate | Sensitivity | Specificity | Precision | F1 Score | R@1 |
| --- | --- | --- | --- | --- | --- | --- | --- | --- | --- | --- |
| 2010 | 160766 | 0.732 | 0.722 | 0.693 | 0.278 | 0.654 | 0.733 | 0.254 | 0.366 | 0.279 |
| 2011 | 231351 | 0.767 | 0.692 | 0.704 | 0.308 | 0.721 | 0.687 | 0.269 | 0.392 | 0.319 |
| 2012 | 330034 | 0.802 | 0.728 | 0.733 | 0.272 | 0.74 | 0.726 | 0.302 | 0.429 | 0.361 |
| 2013 | 468100 | 0.8 | 0.739 | 0.733 | 0.261 | 0.726 | 0.741 | 0.31 | 0.434 | 0.402 |
| 2014 | 648173 | 0.822 | 0.766 | 0.752 | 0.234 | 0.733 | 0.771 | 0.339 | 0.463 | 0.414 |
| 2015 | 858263 | 0.824 | 0.759 | 0.752 | 0.241 | 0.744 | 0.761 | 0.333 | 0.46 | 0.449 |
| 2016 | 1106765 | 0.809 | 0.763 | 0.749 | 0.237 | 0.73 | 0.768 | 0.319 | 0.444 | 0.474 |
| 2017 | 1397964 | 0.783 | 0.757 | 0.732 | 0.243 | 0.698 | 0.767 | 0.31 | 0.428 | 0.483 |
| 2018 | 1734575 | 0.831 | 0.757 | 0.758 | 0.243 | 0.759 | 0.757 | 0.334 | 0.464 | 0.506 |
| 2019 | 2095551 | 0.85 | 0.785 | 0.776 | 0.215 | 0.764 | 0.789 | 0.367 | 0.495 | 0.527 |
| 2020 | 2493887 | 0.82 | 0.782 | 0.766 | 0.218 | 0.743 | 0.788 | 0.342 | 0.468 | 0.525 |
| 2021 | 3014957 | 0.857 | 0.776 | 0.782 | 0.224 | 0.79 | 0.774 | 0.362 | 0.495 | 0.552 |
| 2022 | 3654626 | 0.852 | 0.773 | 0.778 | 0.227 | 0.785 | 0.771 | 0.355 | 0.489 | 0.573 |
| 2023 | 4326353 | 0.865 | 0.792 | 0.79 | 0.208 | 0.787 | 0.793 | 0.38 | 0.512 | 0.587 |

**Supplementary Table 11:**
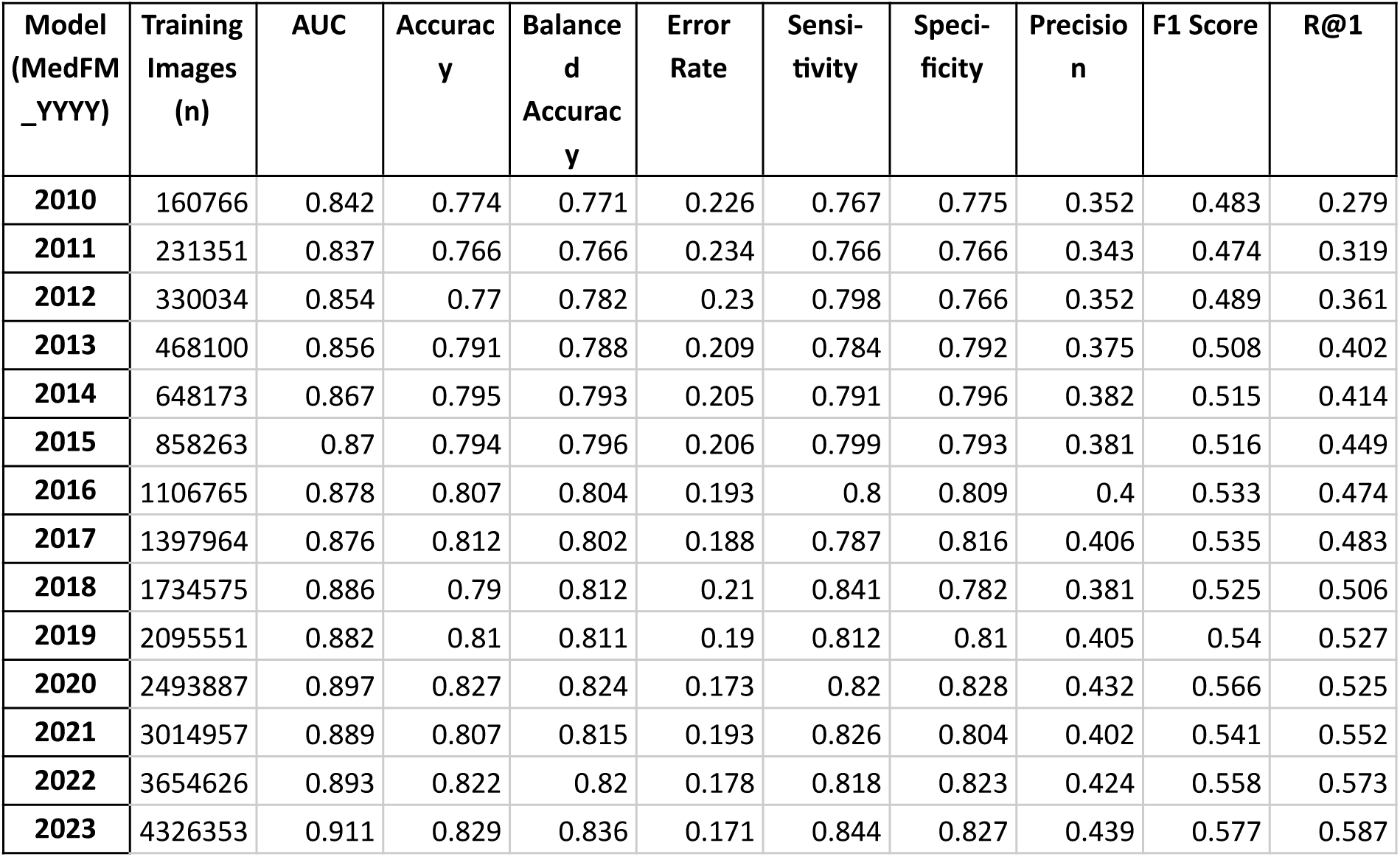
Skin Cancer - All Samples.

| Model<br>(MedFM<br>_YYYY) | Training<br>Images<br>(n) | AUC | Accuracy | Balanced<br>Accuracy | Error<br>Rate | Sensitivity | Specificity | Precision | F1 Score | R@1 |
| --- | --- | --- | --- | --- | --- | --- | --- | --- | --- | --- |
| 2010 | 160766 | 0.842 | 0.774 | 0.771 | 0.226 | 0.767 | 0.775 | 0.352 | 0.483 | 0.279 |
| 2011 | 231351 | 0.837 | 0.766 | 0.766 | 0.234 | 0.766 | 0.766 | 0.343 | 0.474 | 0.319 |
| 2012 | 330034 | 0.854 | 0.77 | 0.782 | 0.23 | 0.798 | 0.766 | 0.352 | 0.489 | 0.361 |
| 2013 | 468100 | 0.856 | 0.791 | 0.788 | 0.209 | 0.784 | 0.792 | 0.375 | 0.508 | 0.402 |
| 2014 | 648173 | 0.867 | 0.795 | 0.793 | 0.205 | 0.791 | 0.796 | 0.382 | 0.515 | 0.414 |
| 2015 | 858263 | 0.87 | 0.794 | 0.796 | 0.206 | 0.799 | 0.793 | 0.381 | 0.516 | 0.449 |
| 2016 | 1106765 | 0.878 | 0.807 | 0.804 | 0.193 | 0.8 | 0.809 | 0.4 | 0.533 | 0.474 |
| 2017 | 1397964 | 0.876 | 0.812 | 0.802 | 0.188 | 0.787 | 0.816 | 0.406 | 0.535 | 0.483 |
| 2018 | 1734575 | 0.886 | 0.79 | 0.812 | 0.21 | 0.841 | 0.782 | 0.381 | 0.525 | 0.506 |
| 2019 | 2095551 | 0.882 | 0.81 | 0.811 | 0.19 | 0.812 | 0.81 | 0.405 | 0.54 | 0.527 |
| 2020 | 2493887 | 0.897 | 0.827 | 0.824 | 0.173 | 0.82 | 0.828 | 0.432 | 0.566 | 0.525 |
| 2021 | 3014957 | 0.889 | 0.807 | 0.815 | 0.193 | 0.826 | 0.804 | 0.402 | 0.541 | 0.552 |
| 2022 | 3654626 | 0.893 | 0.822 | 0.82 | 0.178 | 0.818 | 0.823 | 0.424 | 0.558 | 0.573 |
| 2023 | 4326353 | 0.911 | 0.829 | 0.836 | 0.171 | 0.844 | 0.827 | 0.439 | 0.577 | 0.587 |

**Supplementary Table 12:** Datasets and data sizes used for training and testing in the Monkeypox evaluation use case.

| Category | Sub-category | Publications and social media | NEJ Cohort | Prospective cohort | DermNet NZ | Danderm | Dermis | DermNet | Esteva | Fitzpatrick 17k | Hellenic | PAD UFES | Total |
| --- | --- | --- | --- | --- | --- | --- | --- | --- | --- | --- | --- | --- | --- |
| Excluded |  | 31 | 5 | 0 | 1,973 | 5 | 0 | 0 | 45,440 | 52 | 48 | 0 | 47,554 |
| Included |  | 518 | 87 | 63 | 12,045 | 3,432 | 6,589 | 19,289 | 75,730 | 16,525 | 2,614 | 2,298 | 139,190 |
| Training |  | 518 | 0 | 0 | 12,045 | 0 | 0 | 0 | 0 | 0 | 0 | 0 | 12,563 |
| Testing |  | 0 | 87 | 63 | 0 | 3,432 | 6,589 | 19,289 | 75,730 | 16,525 | 2,614 | 2,298 | 126,627 |
| age | 0-18 y.o. | 14 | 0 | 0 | 0 | 0 | 509 | 0 | 0 | 0 | 0 | 40 | 563 |
|  | 19-64 y.o. | 150 | 0 | 0 | 0 | 0 | 1,932 | 0 | 0 | 0 | 0 | 1,251 | 3,333 |
|  | >=65 y.o | 0 | 0 | 0 | 0 | 0 | 572 | 0 | 0 | 0 | 0 | 1,007 | 1,579 |
|  | Unknown | 354 | 87 | 63 | 12,045 | 3,432 | 3,576 | 19,289 | 75,730 | 16,525 | 2,614 | 0 | 133,715 |
| sex | Female | 26 | 0 | 0 | 0 | 0 | 2,520 | 0 | 0 | 0 | 0 | 753 | 3,299 |
|  | Male | 406 | 87 | 63 | 0 | 0 | 2,593 | 0 | 0 | 0 | 0 | 741 | 3,890 |
|  | Unknown | 86 | 0 | 0 | 12,045 | 3,432 | 1,476 | 19,289 | 75,730 | 16,525 | 2,614 | 804 | 132,001 |
| mpox | yes | 518 | 87 | 63 | 0 | 0 | 0 | 0 | 0 | 0 | 0 | 0 | 668 |
|  | no | 0 | 0 | 0 | 12,045 | 3,432 | 6,589 | 19,289 | 75,730 | 16,525 | 2,614 | 2,298 | 138,522 |
|  | Unknown | 0 | 0 | 0 | 0 | 0 | 0 | 0 | 0 | 0 | 0 | 0 | 0 |
| skin tone (Fitzpatrick) | Fitzpatrick I | 20 | 3 | 0 | 0 | 0 | 0 | 0 | 0 | 2,941 | 0 | 153 | 3,117 |
|  | Fitzpatrick II | 147 | 34 | 26 | 0 | 0 | 0 | 0 | 0 | 4,796 | 0 | 876 | 5,879 |
|  | Fitzpatrick III | 140 | 28 | 27 | 0 | 0 | 0 | 0 | 0 | 3,296 | 0 | 392 | 3,883 |
|  | Fitzpatrick IV | 65 | 15 | 0 | 0 | 0 | 0 | 0 | 0 | 2,775 | 0 | 62 | 2,917 |
|  | Fitzpatrick V | 49 | 6 | 10 | 0 | 0 | 0 | 0 | 0 | 1,527 | 0 | 10 | 1,602 |
|  | Fitzpatrick VI | 97 | 1 | 0 | 0 | 0 | 0 | 0 | 0 | 628 | 0 | 1 | 727 |
|  | Unknown | 0 | 0 | 0 | 12,045 | 3,432 | 6,589 | 19,289 | 75,730 | 562 | 2,614 | 804 | 121,065 |
| body region | Head | 106 | 16 | 2 | 0 | 0 | 1,443 | 0 | 0 | 0 | 0 | 842 | 2,409 |
|  | Neck | 2 | 0 | 1 | 0 | 0 | 96 | 0 | 0 | 0 | 0 | 93 | 192 |
|  | Torso | 75 | 5 | 8 | 0 | 0 | 705 | 0 | 0 | 0 | 0 | 564 | 1,357 |
|  | Upper extremity | 120 | 14 | 26 | 0 | 0 | 916 | 0 | 0 | 0 | 0 | 710 | 1,786 |
|  | Lower extremity | 39 | 9 | 12 | 0 | 0 | 813 | 0 | 0 | 0 | 0 | 89 | 962 |
|  | Anogenital | 100 | 42 | 9 | 0 | 0 | 223 | 0 | 0 | 0 | 0 | 0 | 374 |
|  | Multiple Body Regions | 48 | 0 | 4 | 0 | 0 | 110 | 0 | 0 | 0 | 0 | 0 | 162 |
|  | Unknown | 28 | 1 | 1 | 12,045 | 3,432 | 2,283 | 19,289 | 75,730 | 16,525 | 2,614 | 0 | 131,948 |

**Supplementary Table 13:**
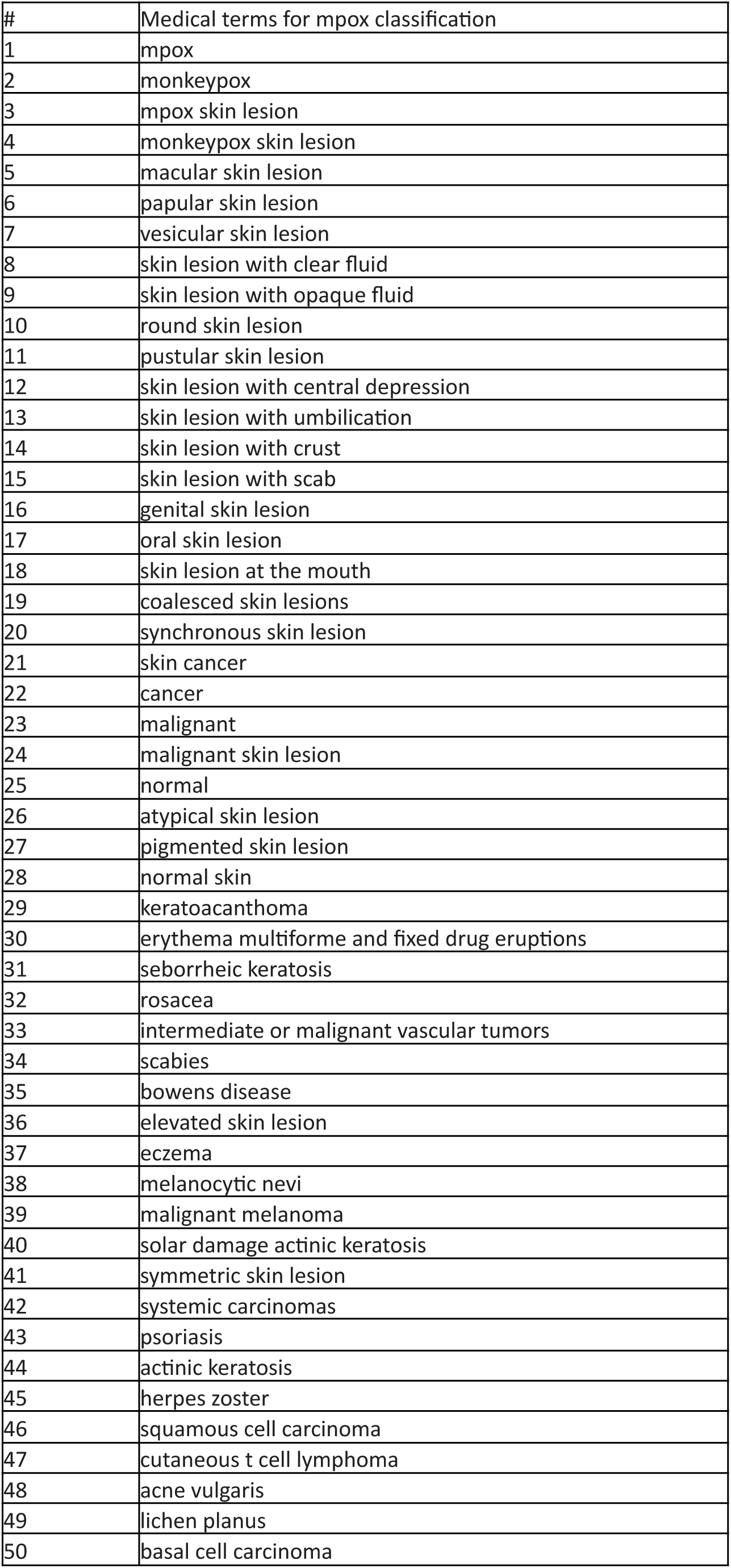
Medical terms for mpox skin lesion classification in clinical photos.

**Supplementary Table 14:** Mpox - Zero-shot.

| Model<br>(MedFM<br>_YYYY) | Training<br>Images<br>(n) | AUC | Accuracy | Balanced<br>Accuracy | Error<br>Rate | Sensitivity | Specificity | Precision | F1<br>Score | R@1 |
| --- | --- | --- | --- | --- | --- | --- | --- | --- | --- | --- |
| 2010 | 160766 | 0.694 | 0.541 | 0.646 | 0.459 | 0.751 | 0.541 | 0.002 | 0.005 | 0.279 |
| 2011 | 231351 | 0.641 | 0.666 | 0.617 | 0.334 | 0.568 | 0.666 | 0.002 | 0.005 | 0.319 |
| 2012 | 330034 | 0.692 | 0.673 | 0.653 | 0.327 | 0.632 | 0.673 | 0.003 | 0.006 | 0.361 |
| 2013 | 468100 | 0.760 | 0.596 | 0.703 | 0.404 | 0.811 | 0.595 | 0.003 | 0.006 | 0.402 |
| 2014 | 648173 | 0.778 | 0.605 | 0.716 | 0.395 | 0.827 | 0.605 | 0.003 | 0.006 | 0.414 |
| 2015 | 858263 | 0.793 | 0.697 | 0.743 | 0.303 | 0.789 | 0.696 | 0.004 | 0.008 | 0.449 |
| 2016 | 1106765 | 0.832 | 0.626 | 0.764 | 0.374 | 0.903 | 0.626 | 0.004 | 0.007 | 0.474 |
| 2017 | 1397964 | 0.831 | 0.698 | 0.765 | 0.302 | 0.832 | 0.698 | 0.004 | 0.008 | 0.483 |
| 2018 | 1734575 | 0.801 | 0.801 | 0.733 | 0.199 | 0.665 | 0.801 | 0.005 | 0.010 | 0.506 |
| 2019 | 2095551 | 0.772 | 0.722 | 0.707 | 0.278 | 0.692 | 0.722 | 0.004 | 0.007 | 0.527 |
| 2020 | 2493887 | 0.802 | 0.720 | 0.735 | 0.280 | 0.751 | 0.719 | 0.004 | 0.008 | 0.525 |
| 2021 | 3014957 | 0.794 | 0.678 | 0.731 | 0.322 | 0.784 | 0.678 | 0.004 | 0.007 | 0.552 |
| 2022 | 3654626 | 0.853 | 0.788 | 0.786 | 0.212 | 0.784 | 0.788 | 0.005 | 0.011 | 0.573 |
| 2023 | 4326353 | 0.834 | 0.744 | 0.766 | 0.256 | 0.789 | 0.744 | 0.004 | 0.009 | 0.587 |

**Supplementary Table 15:** Mpox - 200 Samples.

| Model<br>(MedFM<br>_YYYY) | Training<br>Images<br>(n) | AUC | Accurac<br>y | Balance<br>d<br>Accurac<br>y | Error<br>Rate | Sensi-<br>tivity | Speci-<br>ficity | Precisio<br>n | F1 Score | R@1 |
| --- | --- | --- | --- | --- | --- | --- | --- | --- | --- | --- |
| 2010 | 160766 | 0.772 | 0.698 | 0.716 | 0.302 | 0.735 | 0.698 | 0.004 | 0.007 | 0.279 |
| 2011 | 231351 | 0.748 | 0.683 | 0.702 | 0.317 | 0.721 | 0.683 | 0.004 | 0.007 | 0.319 |
| 2012 | 330034 | 0.794 | 0.700 | 0.727 | 0.300 | 0.755 | 0.699 | 0.004 | 0.007 | 0.361 |
| 2013 | 468100 | 0.829 | 0.727 | 0.764 | 0.273 | 0.800 | 0.727 | 0.004 | 0.009 | 0.402 |
| 2014 | 648173 | 0.826 | 0.741 | 0.761 | 0.259 | 0.781 | 0.741 | 0.005 | 0.009 | 0.414 |
| 2015 | 858263 | 0.822 | 0.760 | 0.758 | 0.240 | 0.757 | 0.760 | 0.005 | 0.009 | 0.449 |
| 2016 | 1106765 | 0.843 | 0.729 | 0.773 | 0.271 | 0.818 | 0.729 | 0.004 | 0.009 | 0.474 |
| 2017 | 1397964 | 0.857 | 0.754 | 0.787 | 0.246 | 0.820 | 0.754 | 0.005 | 0.010 | 0.483 |
| 2018 | 1734575 | 0.826 | 0.737 | 0.777 | 0.263 | 0.817 | 0.737 | 0.005 | 0.010 | 0.506 |
| 2019 | 2095551 | 0.876 | 0.783 | 0.805 | 0.217 | 0.828 | 0.783 | 0.006 | 0.012 | 0.527 |
| 2020 | 2493887 | 0.888 | 0.781 | 0.815 | 0.219 | 0.849 | 0.780 | 0.006 | 0.012 | 0.525 |
| 2021 | 3014957 | 0.918 | 0.839 | 0.851 | 0.161 | 0.863 | 0.839 | 0.008 | 0.016 | 0.552 |
| 2022 | 3654626 | 0.914 | 0.840 | 0.839 | 0.160 | 0.838 | 0.840 | 0.008 | 0.016 | 0.573 |
| 2023 | 4326353 | 0.921 | 0.855 | 0.850 | 0.145 | 0.846 | 0.855 | 0.009 | 0.017 | 0.587 |

**Supplementary Table 16:** Mpox - All Samples.

| Model<br>(MedFM<br>_YYYY) | Training<br>Images<br>(n) | AUC | Accuracy | Balance<br>d<br>Accuracy | Error<br>Rate | Sensi-<br>tivity | Speci-<br>ficity | Precisio<br>n | F1 Score | R@1 |
| --- | --- | --- | --- | --- | --- | --- | --- | --- | --- | --- |
| **2010** | 160766 | 0.852 | 0.806 | 0.776 | 0.194 | 0.746 | 0.806 | 0.006 | 0.011 | 0.279 |
| **2011** | 231351 | 0.840 | 0.795 | 0.784 | 0.205 | 0.773 | 0.795 | 0.005 | 0.011 | 0.319 |
| **2012** | 330034 | 0.864 | 0.857 | 0.782 | 0.143 | 0.708 | 0.857 | 0.007 | 0.014 | 0.361 |
| **2013** | 468100 | 0.886 | 0.770 | 0.815 | 0.230 | 0.859 | 0.770 | 0.005 | 0.011 | 0.402 |
| **2014** | 648173 | 0.878 | 0.811 | 0.816 | 0.189 | 0.822 | 0.811 | 0.006 | 0.013 | 0.414 |
| **2015** | 858263 | 0.878 | 0.814 | 0.791 | 0.186 | 0.768 | 0.815 | 0.006 | 0.012 | 0.449 |
| **2016** | 1106765 | 0.889 | 0.756 | 0.813 | 0.244 | 0.870 | 0.756 | 0.005 | 0.010 | 0.474 |
| **2017** | 1397964 | 0.911 | 0.751 | 0.832 | 0.249 | 0.914 | 0.751 | 0.005 | 0.011 | 0.483 |
| **2018** | 1734575 | 0.919 | 0.866 | 0.852 | 0.134 | 0.838 | 0.866 | 0.009 | 0.018 | 0.506 |
| **2019** | 2095551 | 0.922 | 0.890 | 0.864 | 0.110 | 0.838 | 0.890 | 0.011 | 0.022 | 0.527 |
| **2020** | 2493887 | 0.953 | 0.848 | 0.899 | 0.152 | 0.951 | 0.847 | 0.009 | 0.018 | 0.525 |
| **2021** | 3014957 | 0.946 | 0.840 | 0.874 | 0.160 | 0.908 | 0.839 | 0.008 | 0.016 | 0.552 |
| **2022** | 3654626 | 0.961 | 0.888 | 0.895 | 0.112 | 0.903 | 0.888 | 0.012 | 0.023 | 0.573 |
| **2023** | 4326353 | 0.970 | 0.870 | 0.913 | 0.130 | 0.957 | 0.870 | 0.011 | 0.021 | 0.587 |

**Supplementary Table 17:**
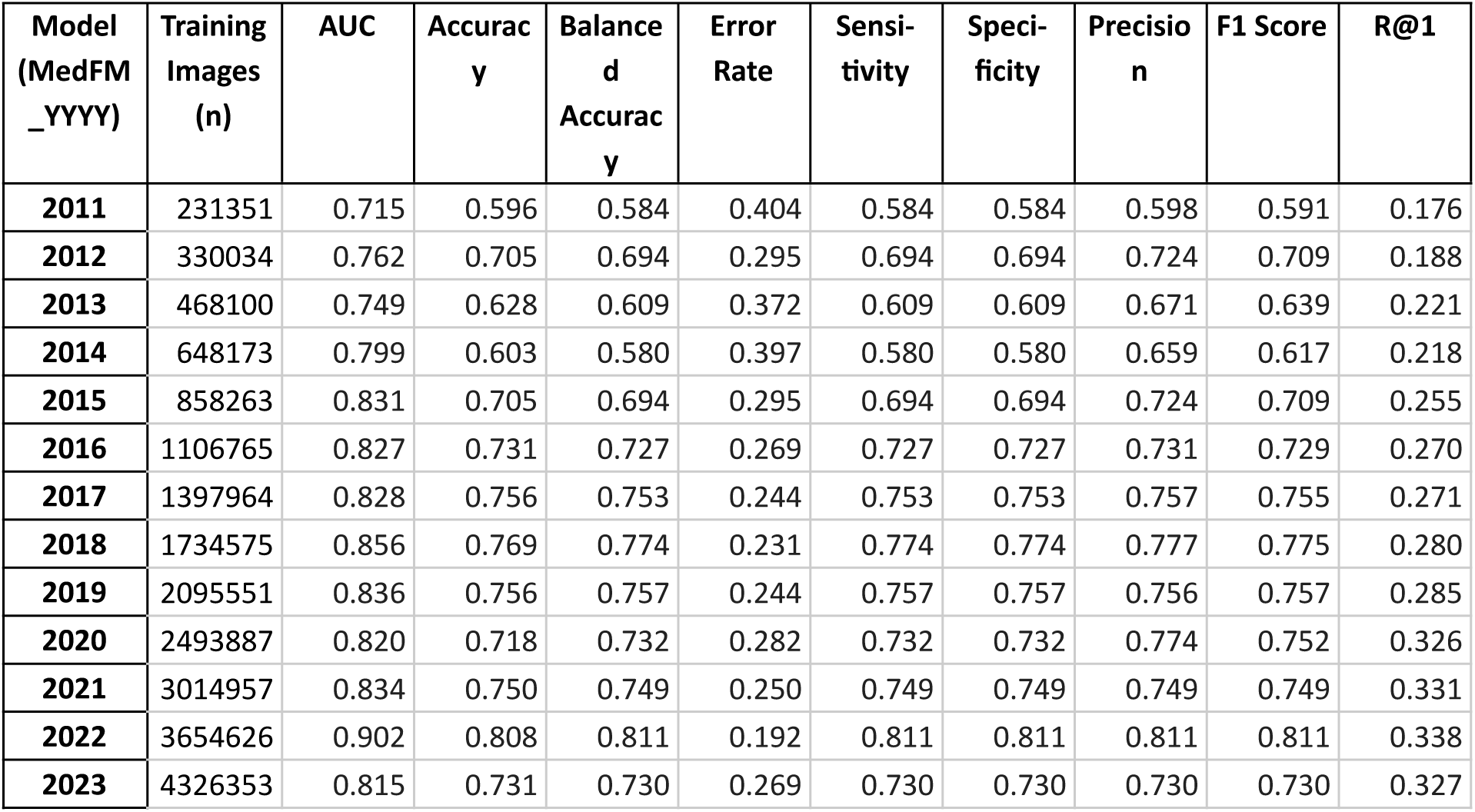
Lung cancer - Zero-shot.

| Model<br>(MedFM<br>_YYYY) | Training<br>Images<br>(n) | AUC | Accuracy | Balance<br>d<br>Accuracy | Error<br>Rate | Sensi-<br>tivity | Speci-<br>ficity | Precisio<br>n | F1 Score | R@1 |
| --- | --- | --- | --- | --- | --- | --- | --- | --- | --- | --- |
| **2011** | 231351 | 0.715 | 0.596 | 0.584 | 0.404 | 0.584 | 0.584 | 0.598 | 0.591 | 0.176 |
| **2012** | 330034 | 0.762 | 0.705 | 0.694 | 0.295 | 0.694 | 0.694 | 0.724 | 0.709 | 0.188 |
| **2013** | 468100 | 0.749 | 0.628 | 0.609 | 0.372 | 0.609 | 0.609 | 0.671 | 0.639 | 0.221 |
| **2014** | 648173 | 0.799 | 0.603 | 0.580 | 0.397 | 0.580 | 0.580 | 0.659 | 0.617 | 0.218 |
| **2015** | 858263 | 0.831 | 0.705 | 0.694 | 0.295 | 0.694 | 0.694 | 0.724 | 0.709 | 0.255 |
| **2016** | 1106765 | 0.827 | 0.731 | 0.727 | 0.269 | 0.727 | 0.727 | 0.731 | 0.729 | 0.270 |
| **2017** | 1397964 | 0.828 | 0.756 | 0.753 | 0.244 | 0.753 | 0.753 | 0.757 | 0.755 | 0.271 |
| **2018** | 1734575 | 0.856 | 0.769 | 0.774 | 0.231 | 0.774 | 0.774 | 0.777 | 0.775 | 0.280 |
| **2019** | 2095551 | 0.836 | 0.756 | 0.757 | 0.244 | 0.757 | 0.757 | 0.756 | 0.757 | 0.285 |
| **2020** | 2493887 | 0.820 | 0.718 | 0.732 | 0.282 | 0.732 | 0.732 | 0.774 | 0.752 | 0.326 |
| **2021** | 3014957 | 0.834 | 0.750 | 0.749 | 0.250 | 0.749 | 0.749 | 0.749 | 0.749 | 0.331 |
| **2022** | 3654626 | 0.902 | 0.808 | 0.811 | 0.192 | 0.811 | 0.811 | 0.811 | 0.811 | 0.338 |
| **2023** | 4326353 | 0.815 | 0.731 | 0.730 | 0.269 | 0.730 | 0.730 | 0.730 | 0.730 | 0.327 |

**Supplementary Table 18:**
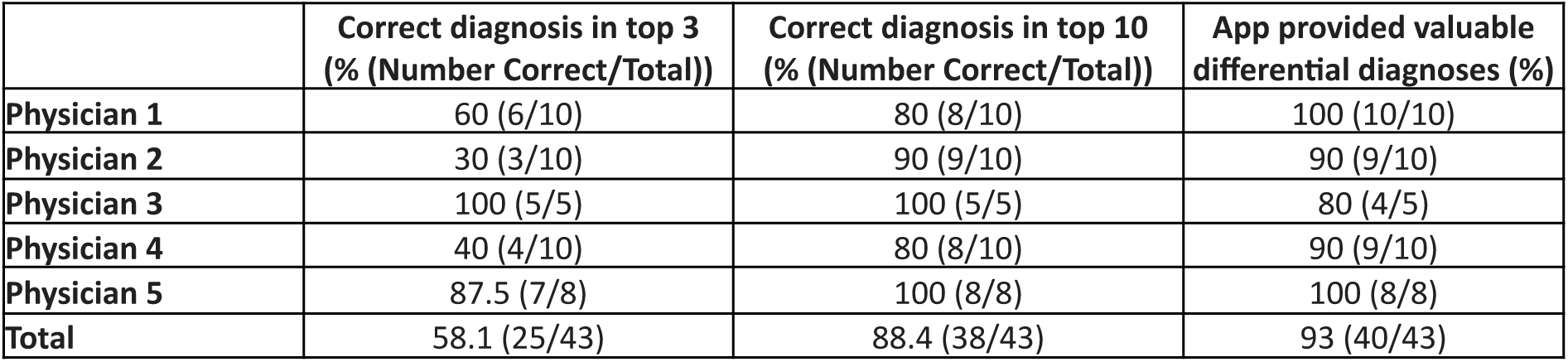
Clinical evaluation of the Image2Paper App.

|  | Correct diagnosis in top 3<br>(% (Number Correct/Total)) | Correct diagnosis in top 10<br>(% (Number Correct/Total)) | App provided valuable<br>differential diagnoses (%) |
| --- | --- | --- | --- |
| Physician 1 | 60 (6/10) | 80 (8/10) | 100 (10/10) |
| Physician 2 | 30 (3/10) | 90 (9/10) | 90 (9/10) |
| Physician 3 | 100 (5/5) | 100 (5/5) | 80 (4/5) |
| Physician 4 | 40 (4/10) | 80 (8/10) | 90 (9/10) |
| Physician 5 | 87.5 (7/8) | 100 (8/8) | 100 (8/8) |
| Total | 58.1 (25/43) | 88.4 (38/43) | 93 (40/43) |

## Supplementary Methods

### Labeling Instructions for MedFM Project

1.) Please only label rows with an empty “investigator” column. Label columns J through X.

2.) Use only the information provided in the sheet. Do not look up the paper.

3.) Primary Goal of this work: Improving the image description.

For each unedited row, the image description initially contains the figure caption. Ensure the description includes:

- What? Diagnosis and procedures (if visible in the image).
- Where? Precise anatomical location of the pathology.
- Who? Relevant patient-specific information (e.g., “patient experienced a car accident” for images with fractures or “patient has a history of diabetes” for images with ulcers). Do not include irrelevant comorbidities. Patient age should generally not be included (except if it is necessary to understand the image). If it is a pediatric case, the information should be included that the patient is a child.

Provide a concise yet comprehensive image description to facilitate the model’s understanding. Remove any unnecessary text such as references, author names, URLs, figure references, dates etc.

4.) **Secondary Goal:** Collecting information on age, sex, skin color, body region, diagnosis, specialty, and modality for sub-cohort analyses.

5.) Use the paper’s terminology for diagnoses. If the image shows an object, use “na” (not applicable). If the image shows complete remission, use “None”

6.) Record skin color only for images displaying skin and not depicting surgical procedures.

For surgical procedures that might involve Betaisodona, affecting skin color, use “u” (unknown).

7.) Sex can be assessed from the text or image if sexual anatomy is visible.

8.) Anatomical regions:

This categorization applies to images with humans. For photos of specimens or histological images use “na” (not applicable).

**Head and neck**

Cephalic (head)

Cervical (neck)

Cranial (skull)

Frontal (forehead)

Nasal (nose)

Occipital (base of skull)

Oral (mouth)

Orbital/ocular (eyes)

**Thorax**

Axillary (armpit)

Costal (ribs)

Deltoid (shoulder)

Mammary (breast)

Pectoral (chest)

Scapular (shoulder blade)

Sternal (breastbone)

Vertebral (backbone)

**Abdomen**

Abdominal (abdomen)

Gluteal (buttocks)

Inguinal (bend of hip)

Lumbar (lower back)

Pelvic (area between hip bones)

Perineal (area between anus and external genitalia)

Pubic (genitals)

Sacral (end of vertebral column)

**Upper extremity**

Antebrachial (forearm)

Antecubital (inner elbow)

Brachial (upper arm)

Carpal (wrist)

Cubital (elbow)

Digital (fingers/toes)

Manual (hand)

Palmar (palm)

**Lower extremity**

Crural (shin, front of lower leg)

Femoral (thigh)

Patellar (front of knee)

Pedal (foot)

Plantar (arch of foot)

Popliteal (back of knee)

Sural (calf, back of lower leg)

Tarsal (ankle)

9.) Please categorize into the following modalities:

**X- ray**

- Plain X-rays
- Panoramic dental X-ray/Orthopantomograph
- Fluoroscopy
- Cephalogram

**Contrast-enhanced radiography**

- Angiography
- Venography
- Cholangiography
- Fistulography
- Hysterosalpingography
- Urography
- Barium swallow (esophagram)
- Upper gastrointestinal series (UGI)
- Barium enema (lower gastrointestinal series)
- Arthrography
- Myelography
- Digital subtraction angiography (DSA)
- Coronary angiogram
- Other contrast-enhanced X-ray techniques

**CT**

- Non-contrast CT
- Contrast-enhanced CT
- CT angiography
- Cone-Beam CT

**MRI**

- Non-contrast MRI
- Contrast-enhanced MRI
- Magnetic Resonance Angiography (MRA)

**Microscopy**

- Brightfield microscopy

- Hematoxylin and eosin (H&E)
- Gram staining
- Acid-fast staining (e.g., Ziehl-Neelsen)
- Giemsa staining
- Silver staining (e.g., Gomori methenamine silver, Warthin-Starry)
- Periodic acid-Schiff (PAS)
- Trichrome staining (e.g., Masson’s trichrome)
- Immunohistochemistry (IHC)
- Phase-contrast microscopy
- Darkfield microscopy
- Polarized light microscopy
- Fluorescence microscopy
- Fluorescence in situ hybridization (FISH)
- Transmission electron microscopy (TEM)

**Ultrasound**

- General ultrasound
- Doppler ultrasound (vascular studies)
- Echocardiography (heart studies)
- Eye and Orbital ultrasound

**Nuclear**

- Positron Emission Tomography (PET)
- Single-Photon Emission Computed Tomography (SPECT)
- Scintigraphy
- Whole Body Bone Scan (Tc99m)

**Endoscopy**

- Colonoscopy
- Laparoscopy
- Arthroscopy
- Esophagoscopy
- Otoscope
- Cystoscopy
- Cervicoscopy
- Colposcopy
- Hysteroscopy
- Thoratocopy
- Laryngoscopy
- Nasopharyngolaryngoscopy
- Esophagogastroduodenoscopy

**OCT**

- Retina and optic nerve
- Anterior segment of the eye

**Funduscopy**

- Plain Fundoscopy
- fluorescein angiography” or “fundus fluorescein angiography” (FFA)

**Dermoscopy Photo**

- Clinical images with smartphones/cameras

Others (less common or specialized imaging techniques)

10.) For the diagnosis, please exactly use the wording of the paper (e.g. after you have identified the diagnosis which can be seen on the image, copypaste it to the diagnosis column).

11.) For the column “Can the diagnosis be seen on the image? (yes/no)”, please answer with no, e.g. if the image shows surgery preparations, healed wounds, or complete remission of cancer, and yes otherwise.

12.) For specialty choose the one that you feel is most appropriate to the clinical situation that is shown in the image, e.g. for a multidisciplinary case involving different specialities, please label the specialty that is leading the clinical situation.

13.) Write out abbreviations in full.

14.) The first 1000 rows have already been filled-in and can be used as examples.

15.) If you encounter any issue in the sheet, e.g. a low quality photo, please use the comment function of Google sheet to mark this row.

### Image Exclusion Criteria

Exclude images if they meet the following criteria:

- Related to veterinary medicine.
- Panel or multiple images.
- Abstract drawings or graphs.
- Any image that is not linked to a human disease, e.g. a group photo of physicians
- figure caption is include in the image
- non-english texts

To exclude an image, write “Exclude” in the Image Description field. Do not exclude an image if:

- It features an animal relevant to human disease (e.g., as a vector).

